# The Frequency, Penetrance and Variable Expressivity of Dilated Cardiomyopathy-Associated Putative Pathogenic Gene Variants in UK Biobank Participants

**DOI:** 10.1101/2021.11.22.21266651

**Authors:** Ravi Shah, Babken Asatryan, Ghaith Sharaf Dabbagh, Nay Aung, Mohammed Y Khanji, Luis R. Lopes, Stefan van Duijvenboden, Anthony Holmes, Daniele Muser, Andrew P. Landstrom, Aaron Mark Lee, Pankaj Arora, Christopher Semsarian, Virend K. Somers, Anjali Owens, Patricia B Munroe, Steffen E. Petersen, C. Anwar A. Chahal

## Abstract

**Background:** There is a paucity of data regarding the phenotype of dilated cardiomyopathy (DCM) gene variants in the general population. We aimed to determine the frequency and penetrance of DCM-associated putative pathogenic gene variants in a general, adult population, with a focus on the expression of clinical and subclinical phenotype, including structural, functional and arrhythmic disease features.

**Methods:** UK Biobank participants who had undergone whole exome sequencing (WES), ECG and cardiovascular magnetic resonance (CMR) imaging were selected for study. Three different variant calling strategies (one primary and two secondary) were used to identify subjects with putative pathogenic variants in 44 DCM genes. The observed phenotype was graded to either 1) DCM (clinical or CMR diagnosis); 2) early DCM features, including arrhythmia and/or conduction disease, isolated ventricular dilation, and hypokinetic non-dilated cardiomyopathy; or 3) phenotype-negative.

**Results:** Among 18,665 individuals included in the study, 1,463 (7.8%) subjects possessed ≥1 putative pathogenic variant in 44 DCM genes by the main variant calling strategy. A clinical diagnosis of DCM was present in 0.34% and early DCM features in 5.7% of individuals with putative pathogenic variants. ECG and CMR analysis revealed evidence of subclinical DCM in an additional 1.6% and early DCM features in 15.9% of individuals with putative pathogenic variants. Arrhythmias and/or conduction disease (15.2%) were the most common early DCM features, followed by hypokinetic non-dilated cardiomyopathy (4%). The combined clinical/subclinical penetrance was ≤30% with all three variant filtering strategies. Clinical DCM was slightly more prevalent among participants with putative pathogenic variants in definitive/strong evidence genes, as compared to those with variants in moderate/limited evidence genes.

**Conclusions:** In the UK Biobank, approximately 1/6 of adults with putative pathogenic variants in DCM genes exhibited a subclinical phenotype based on ECG and/or CMR, most commonly manifesting with arrhythmias in the absence of substantial ventricular dilation/dysfunction.

**Clinical Perspective:** *What is new?:*

- Among individuals with putative pathogenic DCM gene variants, subclinical DCM and early DCM features, detected by ECG and/or CMR, were nearly four times more common than clinically manifest DCM or early features (23.7% vs. 6.1%).
- Over 90% of subjects with a putative pathogenic variant in DCM-associated genes did not have a prior history of DCM.
- Clinical DCM was slightly more prevalent among participants with putative pathogenic variants in definitive/strong evidence genes (13.9% for clinical and subclinical), as compared to those with variants in moderate/limited evidence genes, but there was no significant difference in combined clinical/subclinical phenotype by cluster.
- The overall clinical/subclinical penetrance of DCM-associated single putative pathogenic variants was highly variable between genes, ranging from 0 to 66.7%.

*What are the clinical implications?:*

- Arrhythmias and cardiac conduction disease are the most common early manifestation of putative pathogenic variants implicated in DCM, mostly occurring prior to the development of structural/functional abnormalities.
- A genotype-first screening approach for DCM using a large genetic panel is currently not suitable in the general population due to incomplete understanding of DCM genetic architecture and reduced penetrance of DCM-associated putative pathogenic variants.

**Journal Subject Terms:** Cardiomyopathy; Genetics; Sudden Cardiac Death

## 1. INTRODUCTION

Dilated cardiomyopathy (DCM) is a genetic heart disease that frequently leads to end-stage heart failure, characterized by progressive left ventricular (LV) or biventricular dilation and impaired contraction that is not explained exclusively by abnormal loading conditions (hypertension or valvular heart disease) or coronary artery disease.1 Patients with DCM often present in adulthood and are prone to life-threatening ventricular arrhythmias, with 30% dying suddenly.2 With the advent of next-generation sequencing technologies, there has been a dramatic increase in the number of genes tested and variants identified in patients with DCM.3 To date, over 250 genes from ten gene ontologies have been reported in association with DCM, of which only 19 were recently found to have moderate, strong, or definitive evidence for causality in monogenic DCM by Clinical Genome Resource (ClinGen) DCM Gene Curation Expert Panel.4 Currently, it is estimated that a pathogenic or likely pathogenic variant can be identified in approximately 20-35% of patients with DCM.5, 6

The increasing availability, falling costs, and widespread use of genetic testing (including direct-to-consumer testing) offer an opportunity to use a ‘genome-first’ method for diagnosis.7 However, routine genetic screening is not currently justified because of the unknown frequency of putative pathogenic DCM gene variants in the general population, as well as uncertainties with incomplete penetrance and variable expressivity, and challenges in variant calling.8, 9 These factors complicate the applicability and clinical implications of a given gene variant. Understanding the frequency and penetrance of DCM-associated gene variants in the general population is critical to patient and family counseling and clinical decision-making in those with incidental findings. However, to date, the prevalence and penetrance of DCM-associated pathogenic variants in the general population remain insufficiently investigated.

Using the UK Biobank, we aimed to: 1) determine the frequency of putative pathogenic variants in the ClinGen DCM Gene Curation Expert Panel (GCEP)-asserted genes;4 2) determine clinical DCM penetrance based on electronic health records; 3) identify patients with subclinical DCM or DCM features using advanced, quantitative 12-lead electrocardiographic (ECG) and cardiovascular magnetic resonance (CMR) imaging data; and 4) assess the impact of putative pathogenic variants in DCM-associated genes on patient outcomes. This study provides a large-scale genotype-phenotype correlation for DCM genes in the middle to older aged adult population and in a subset of participants with clinically diagnosed DCM, with a focus on the expression of clinical and subclinical phenotype, and considering structural and arrhythmic features of DCM.

## 2. METHODS

### 2.1. Study population

The UK Biobank study is a prospective study of 502,493 UK residents aged between 40-69 years at enrollment, who were recruited at 22 assessment centers across the United Kingdom.10 Participants attended a center visit undergoing deep phenotyping, including anthropometric measurements, extensive health and lifestyle questionnaires and biological samples. This provided information on baseline characteristics and self-reported medical conditions. Additional links to primary care records and external hospital data records provided, in the form of ICD-10 diagnostic codes and OPCS-4 operation codes, data from hospital admissions. The survival status was updated until January 2018, generating long-term follow-up data. A subset of participants in the UK Biobank have undergone a selection of whole exome sequencing (WES), CMR and 12-lead ECG recordings; this subset comprised the cohort of this study. The UK Biobank received approval from the North West Multi-Centre Research Ethics Committee.

### 2.2. Gene-first approach to identify the study population

Amongst UK Biobank participants, 200,000 underwent WES as previously described.11 For this study, we used a panel of 44 genes recently asserted to be implicated in DCM by the ClinGen DCM GCEP.4 This panel includes 11 genes with definite evidence (*BAG3, DES, FLNC, LMNA, MYH7, PLN, RBM20, SCN5A, TNNC1, TNNT2, TTN*), one with strong evidence (*DSP*), seven with moderate evidence (*ACTC1, ACTN2, JPH2, NEXN, TNNI3, TPM1, VCL*) and 25 with limited evidence for causality in monogenic DCM (*ABCC9, ANKRD1, CSRP3, CTF1, DSG2, DTNA, EYA4, GATAD1, ILK, LAMA4, LDB3, MYBPC3, MYH6, MYL2, MYPN, NEBL, NKX2-5, OBSCN, PLEKHM2, PRDM16, PSEN2, SGCD, TBX20, TCAP, TNNI3K*).4 We used three variant filtering strategies (one primary and two secondary) to classify variants. For all strategies, we restricted the analysis to only high quality (read depth ≥10, call quality ≥20, and genotype quality ≥20) and rare variants (minor allele frequency ≤0.001 in both gnomAD12 and the UK Biobank exome dataset). A separate analysis was performed for the American College of Medical Genetics and Genomics (ACMG) clinically actionable DCM genes (*TNNT2, LMNA, FLNC*, and *TTN)*.13

In the first filtering strategy (“missense pLOF FAF”, main strategy), we used ANNOVAR14 annotations and REVEL scores (a method for predicting deleterious missense variants15) to determine a set of putative pathogenic variants (as used elsewhere16, 17). Variants with ANNOVAR annotations of either frameshift insertions/deletions, gain/loss of stop codon, or disruption of canonical splice site dinucleotides were classified as predicted loss-of-function variant (pLOF). Missense variants were determined as predicted pathogenic if the annotated REVEL score was ≥0.65.16 For *TTN*, only radical variants (i.e., nonsense, frameshift, and splice-site variants) were considered. We applied a further filtering allele frequency (FAF), removing all variants with a FAF of 8.4 × 10^−5^ or greater in gnomAD and/or UK Biobank18 to produce our final set of variants. Due to the population prevalence of DCM, variants that occur more frequently than this are unlikely to be causative variants under a monogenic Mendelian model. This frequency threshold for DCM and other inherited cardiac conditions has been previously defined.18

Two secondary variant filtering strategies were performed (“InterVar FAF” and “InterVar FAF ClinVar”). Criteria used for these variant filtering strategies are provided in the **Supplementary Methods**.

### 2.3. Quality control of variant filtering strategy based on clinical DCM population

Prior to applying our genetic testing approach to the study population, we performed a quality-control analysis of the filtering and variant calling strategies on individuals with the clinical diagnosis of DCM WES (see subsection 3.1 in the Results).

### 2.4. ECG analysis

All individuals who underwent CMR also underwent 12-ECG recording. Briefly, 10 electrodes were placed in standard position, recorded at a frequency of 500 Hz for 10 seconds (Cardiosoft v6.51 GE, Wauwatosa, WI, USA) and stored in XML file format. These files were downloaded and reprocessed using GE MUSE v9.0 SP4, Marquette 12 SL.19 Unusable ECG tracings were manually confirmed and removed. Of the remaining, 100 were randomly selected and underwent manual review by a board-certified cardiologist masked to the clinical diagnoses, CMR or genetic status. These were then classified into brady- and tachyarrhythmias, conduction system disease using established criteria (see **Table S1** for details).20

### 2.5. CMR analysis

The UK Biobank CMR protocol has been described previously.21 In brief, all CMR scans were acquired on a wide-bore 1.5 Tesla scanner (MAGNETOM Aera, Syngo Platform VD13A, Siemens Healthcare, Erlangen, Germany). The practical and ethical considerations posed by the large scale and observational nature of the UKB preclude the use of contrast or stress agents. The protocol includes bright blood anatomic assessment (sagittal, coronal, and axial), balanced and steady-state free precession (SSFP) sequences, left and right ventricular SSFP cine images (long and short axes), myocardial tagging (three short-axis slices), native T1 mapping, aortic flow, and imaging of the thoracic aorta. Typical parameters were: TR/TE = 2.6/1.1 ms, flip angle 80°, GRAPPA factor 2, voxel size 1.8 × 1.8 × 8 mm^3^ (6 mm for long axis). The actual temporal resolution of 32 ms was interpolated to 50 phases per cardiac cycle (∼20 ms). Analysis was performed using Circle CVI post-processing software (Version 5.1.1, Circle Cardiovascular Imaging Inc., Calgary, Canada).22 Further details on phenotyping are given in the **Supplementary Appendix**.

### 2.6. Penetrance analysis

We defined penetrant disease based on the DCM clinical spectrum as laid out in the 2016 ESC position statement on Dilated Cardiomyopathy.23 Briefly, the spectrum includes DCM (LV dilation and hypokinesia), hypokinetic non-dilated cardiomyopathy (hypokinesia without LV dilatation), isolated LV dilation (LV dilation without hypokinesia) or arrhythmia and/or conduction disturbances.23

Phenotypic definitions were based on a combination of clinical diagnosis (self-reported conditions and ICD-10 codes), procedures (self-reported and OPCS-4 codes), 12-ECGs, and cardiac CMR imaging (where available). A full list of phenotype definitions is shown in **Table S1** and is adapted from definitions used elsewhere.24-26 The observed phenotype was graded to either 1) clinically diagnosed DCM; 2) early DCM features, including arrhythmia and/or cardiac conduction disease (CCD), isolated ventricular dilation, and subclinical DCM; or 3) phenotype-negative. Clinical DCM was defined by the presence of ICD-10 I42.0 whereas subclinical DCM was defined by the fulfillment of the CMR criteria for DCM in the absence of a clinical history of DCM. In the classification of phenotype, the prioritization of phenotype categories was as follows: clinical DCM > subclinical DCM > hypokinetic non-dilated cardiomyopathy > isolated ventricular dilatation > arrhythmia and/or CCD. For example, in the presence of ICD-10 code I42.0, the subject was considered to have clinical DCM regardless of other history features and ECG/CMR features. The diagnosis of hypokinetic non-dilated cardiomyopathy, isolated ventricular dilatation, and subclinical DCM derived from analysis of CMR data. The phenotype category ‘arrhythmia and/or CCD’ was defined as atrial fibrillation/flutter, bradyarrhythmia, CCD, pre-excitation syndrome, or ventricular arrhythmia.

The penetrance and outcome analyses was stratified based on gene-evidence clusters as defined by the ClinGen DCM Gene Curation Expert Panel.4 Accordingly, genes were clustered in the following categories: definitive/strong, moderate, or limited evidence.

### 2.7. Analysis of genetic yield in the clinical DCM population (Quality Control)

Patients with a clinical DCM were identified from the UK Biobank population using ICD-10 code I42.0. Patients without clinically significant coronary artery disease were included; those with myocardial infarction or revascularization26, were excluded. Genetic yield was determined for ClinGen DCM GCEP-asserted DCM associated genes and classified according to evidence category,4 using the same filtering strategies as described above.

### 2.8. Statistical analysis

Statistical analysis was performed with R statistical computing and graphics software, Version 3.6.1,27 using *tidyverse*28 and *tableone*29 packages. Continuous, normally distributed data is summarized as mean (standard deviation) and non-normally distributed data as median (interquartile range [IQR]). Continuous data were compared using a two-sample *t*-test and categorical data using a Chi-squared test to test for differences between genotype-positive and genotype-negative individuals. Details regarding outcome analysis are provided in the **Supplementary Methods**.

## 3. RESULTS

### 3.1. Quality control of variant filtering strategy based on clinical DCM population

Among 502,462 UK Biobank participants, there were 1415 (0.28%) individuals with the known clinical diagnosis of non-ischemic DCM (30.2% female, mean age 59.8±7 years at enrolment). **Table S2** shows the demographic characteristics of these patients. Among DCM patients, 340 (24%) individuals underwent WES. Screening of genes ascertained to have at least limited evidence for causality in monogenic DCM revealed putative pathogenic variants in 55 (16%) subjects (**Figure S1**). In accordance with previous observations,30-32 truncating variants in the *TTN* gene were the most common (n=17, 31% of genotype-positive DCM cases, 5% of all genotyped DCM cases), followed by *DSP* variants >1 putative pathogenic variants (n=5 for each, 9.1% of genotype-positive DCM cases, 1.5% of all genotyped DCM cases). These observations validate our primary variant filtering strategy as one in line with that applied in clinical practice. Genetic test results in the clinical DCM subset using secondary variant filtering strategies are summarized in **Figure S1**.

### 3.2. Study population

Out of 502,462 participants in the UK Biobank (54.4% female), 200,619 had undergone WES; 42,078 had 12-ECG, 39,616 had CMR. Given the staged approach to participant accrual, 18,665 participants had WES, 12-ECG, and CMR forming the study population (52.7% female; average age: 55 years at recruitment and 64.4 years at last follow-up; Figure 1). Arrhythmia and/or cardiac conduction disease was present in 2,729 (14.6%), isolated ventricular dilation in 522 (2.8%), hypokinetic non-dilated cardiomyopathy in 645 (3.5%), and clinical/subclinical DCM in 189 (1%) subjects (Table 1).

**Table 1.** Demographic and clinical characteristics of the study population and of phenotypic subgroups.

|  | **Overall** | **Phenotype negative** | **Arrhythmia and/or CCD** | **Isolated ventricular dilatation** | **Hypokinetic non-dilated cardiomyopathy** | **DCM\*** |
| --- | --- | --- | --- | --- | --- | --- |
| **N** | 18665 | 14580 | 2729 | 522 | 645 | 189 |
| **Female, n (%)** | 9844 (52.7) | 8345 (57.2) | 938 (34.4) | 281 (53.8) | 195 (30.2) | 85 (45.0) |
| **Age at recruitment, yrs** | 54.96 (7.50) | 54.33 (7.44) | 57.33 (7.31) | 55.45 (6.86) | 57.62 (7.33) | 58.70 (6.98) |
| **Age at last follow up, yrs** | 64.38 (7.47) | 63.78 (7.41) | 66.72 (7.32) | 64.82 (6.81) | 66.96 (7.25) | 67.99 (6.96) |
| **Systolic BP automated reading, mmHg, mean (SD)** | 136.75 (18.66) | 135.84 (18.37) | 140.70 (19.33) | 140.12 (19.75) | 136.80 (18.27) | 140.23 (20.10) |
| **Diastolic BP automated reading, mmHg, mean (SD)** | 81.25 (10.37) | 81.07 (10.27) | 82.36 (10.63) | 79.26 (10.59) | 82.34 (10.76) | 81.20 (11.24) |
| **Body mass index, kg/m2** | 26.51 (4.16) | 26.43 (4.13) | 26.89 (4.21) | 25.90 (4.20) | 27.06 (4.30) | 27.09 (4.39) |
| **Body surface area, m2** | 1.86 (0.21) | 1.85 (0.20) | 1.92 (0.21) | 1.83 (0.19) | 1.93 (0.22) | 1.88 (0.21) |
| **Creatinine, (μmol/L)** | 72.12 (14.09) | 71.34 (13.88) | 75.68 (14.67) | 70.44 (13.25) | 76.46 (14.30) | 71.92 (13.00) |
| **Dead, n (%)** | 221 (1.2) | 131 (0.9) | 61 (2.2) | 8 (1.5) | 16 (2.5) | 5 (2.6) |
| **Age at death, yrs** | 70.73 (6.71) | 69.34 (6.94) | 72.32 (5.66) | 72.12 (6.71) | 74.36 (6.57) | 73.60 (4.56) |
| **Coronary artery disease, n (%)** | 846 (4.5) | 429 (2.9) | 281 (10.3) | 32 (6.1) | 72 (11.2) | 32 (16.9) |
| **Heart failure, n (%)** | 240 (1.3) | 60 (0.4) | 96 (3.5) | 20 (3.8) | 40 (6.2) | 24 (12.7) |
| **Hypertension, n (%)** | 6028 (32.3) | 4324 (29.7) | 1192 (43.7) | 168 (32.2) | 254 (39.4) | 90 (47.6) |
| **Stroke, n (%)** | 330 (1.8) | 213 (1.5) | 88 (3.2) | 13 (2.5) | 13 (2.0) | 3 (1.6) |
| **Valvular heart disease, n (%)** | 527 (2.8) | 229 (1.6) | 197 (7.2) | 43 (8.2) | 45 (7.0) | 13 (6.9) |
| **Atrial fibrillation/flutter, n (%)** | 832 (4.5) | 0 (0.0) | 646 (23.7) | 41 (7.9) | 124 (19.2) | 21 (11.1) |
| **Bradyarrhythmia, n (%)** | 352 (1.9) | 0 (0.0) | 273 (10.0) | 26 (5.0) | 31 (4.8) | 22 (11.6) |
| **Cardiac conduction defect, n (%)** | 2448 (13.1) | 0 (0.0) | 2141 (78.5) | 125 (23.9) | 123 (19.1) | 59 (31.2) |
| **Sick sinus syndrome, n (%)** | 19 (0.1) | 0 (0.0) | 13 (0.5) | 5 (1.0) | 1 (0.2) | 0 (0.0) |
| **CIED, n (%)** | 148 (0.8) | 0 (0.0) | 120 (4.4) | 16 (3.1) | 8 (1.2) | 4 (2.1) |
| **Pre-excitation syndrome, n (%)** | 35 (0.2) | 0 (0.0) | 32 (1.2) | 1 (0.2) | 2 (0.3) | 0 (0.0) |
| **Ventricular arrhythmia, n (%)** | 69 (0.4) | 0 (0.0) | 56 (2.1) | 4 (0.8) | 6 (0.9) | 3 (1.6) |

BP = blood pressure; DCM = dilated cardiomyopathy; CIED = cardiac implantable electronic device (including single- and dual-chamber permanent pacemaker, implantable cardioverter defibrillator, cardiac resynchronization therapy). \*DCM determined by MRI data only.

**Figure 1.**
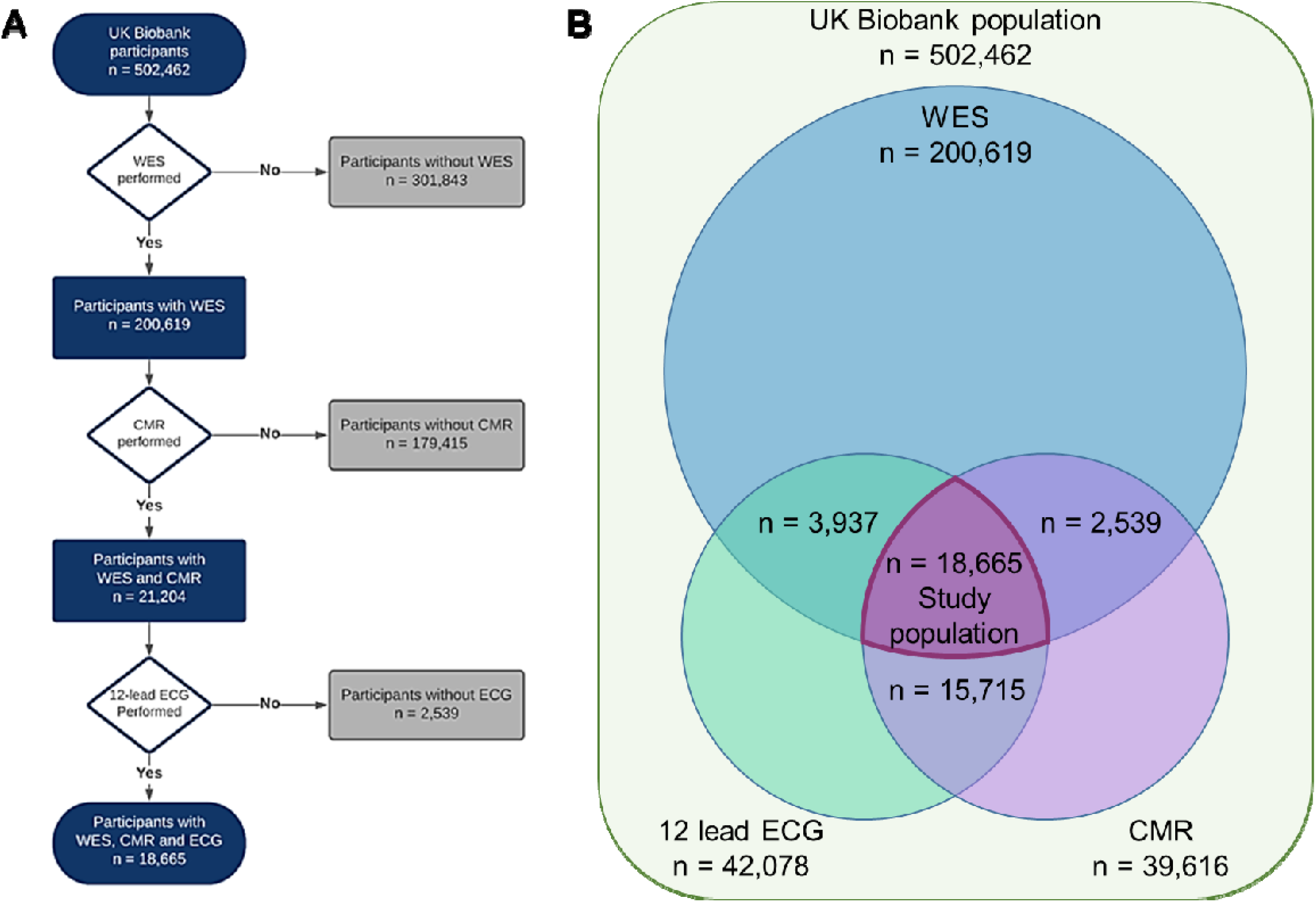
Study population selection criteria. A, Flowchart demonstrates the sequential inclusion/exclusion criteria for the study population. B, Venn diagram showing the number of participants within the whole UK Biobank population with whole exome sequencing (WES), 12-lead ECG, and cardiac magnetic resonance (CMR) imaging.

### 3.3. Prevalence of DCM-associated putative pathogenic variants in the UK Biobank

Among 18,665 individuals, 1,463 (7.8%) were found to host at least one putative pathogenic variant in DCM-associated genes using the primary variant filtering strategy (Figure 2). Putative pathogenic variants were found in all 44 screened genes, and most frequently affected *OBSCN* (n=153, 10.5% of all DCM genotype-positives), *MYH6* (n=149, 10.2%), *SCN5A* (n=140, 9.6%), *MYH7* (n=122, 8.3%), *FLNC* (n=121, 8.3%), *MYBPC3* (n=46, 3.1%), and *TTN* genes (n=44, 3%). There were 30 individuals with *LMNA* variants (2%). Sixty-five individuals (4.4%) carried two or more putative pathogenic variants in the same or different genes. The prevalence of putative pathogenic variants according to secondary filtering strategies are provided in Supplementary Material (**Figure S2**).

**Figure 2.**
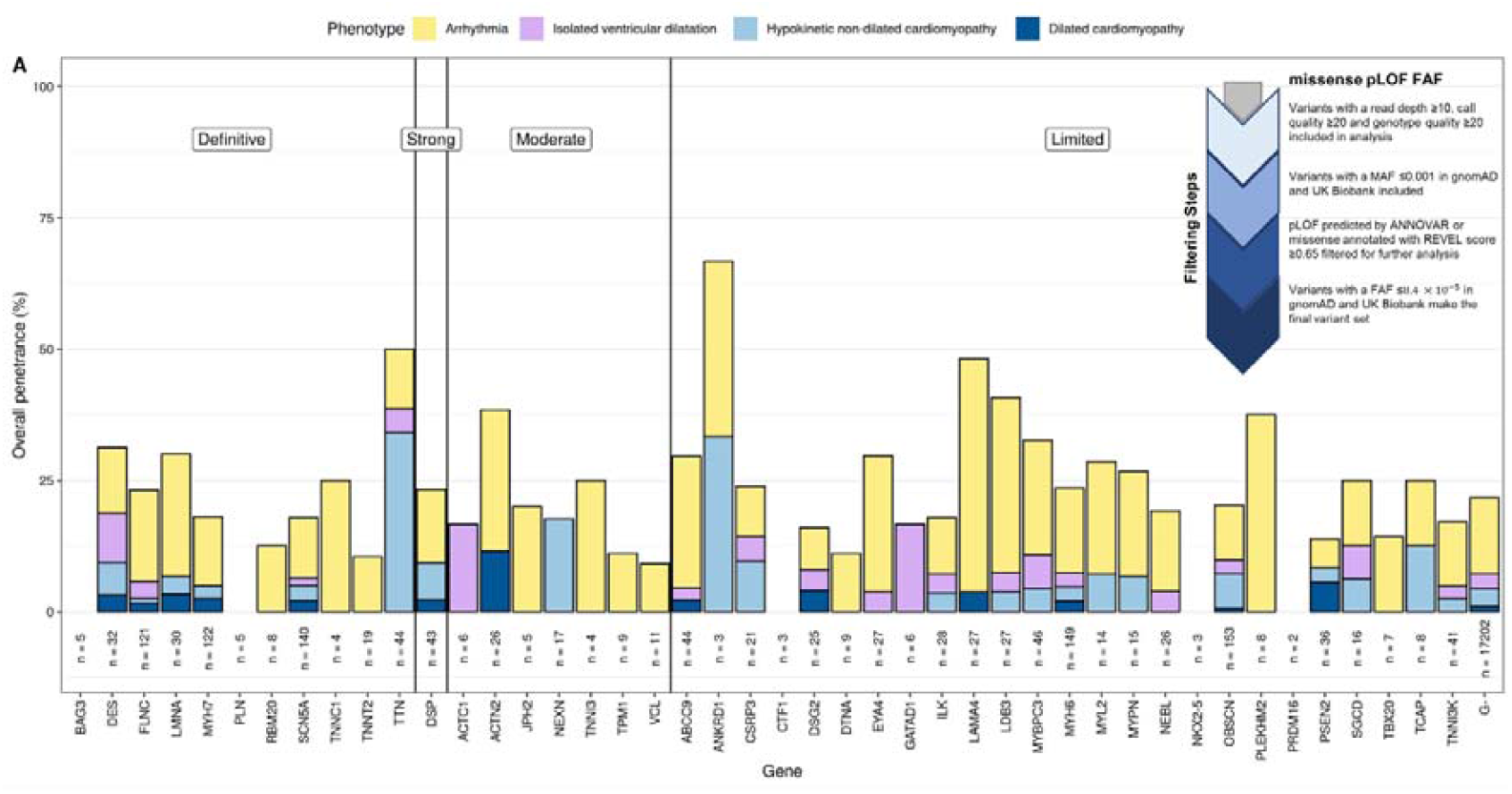
Clinical and subclinical penetrance of putative pathogenic variants DCM-associated genes in middle and older aged adults. For each DCM-associated gene, the height of the bar indicates the percentage of pathogenic/likely pathogenic variant carriers, by missense pLOF FAF filtering strategy, with the specified phenotypes. Total number of participants with a pathogenic/likely pathogenic variant for each DCM-associated gene is indicated below the bar. The phenotype prevalence in those without a pathogenic or likely pathogenic mutation is shown on the far right (labelled “G–“). Genes are categorized according to the strength of evidence determined by the ClinGen DCM gene curation expert panel and ordered alphabetically within each category.

### 3.4. Clinical disease penetrance of DCM-associated putative pathogenic variants in the UK Biobank

Among 1,463 putative pathogenic variant carriers, five individuals had a clinical diagnosis of DCM; 14 additional individuals diagnosed with DCM did not host any putative pathogenic variants (Table 2). Those with putative pathogenic variants more frequently had heart failure (2.1 vs. 1.2%, p=0.01), but the risk of developing heart failure was not different between groups (HR 1.46 [CI 0.96 – 2.24]). Patients with putative pathogenic variants did not show any difference in LVEF, LVESV, LVEDV, age at recruitment, death at follow-up, or age at death, in comparison to those without (Table 3). The comparison of demographic and clinical characteristics of participants with and without putative pathogenic variants according to secondary variant filtering strategies is provided in **Table S3**.

**Table 2.** Clinical and subclinical penetrance of DCM and DCM-associated clinical features in participants with putative pathogenic DCM variants.

|  | Filter | Overall | Phenotype negative | Phenotype positive |  |  |  |
| --- | --- | --- | --- | --- | --- | --- | --- |
|  |  |  |  | Early DCM features |  |  | DCM |
|  |  |  |  | Arrhythmia and/or cardiac conduction disease | Isolated ventricular dilatation | Hypokinetic non-dilated cardiomyopathy |  |
| **Clinical phenotype** | Total | 18665 | 17545 (94%) | 1101 (5.9%) | NA | NA | 19 (0.1%) |
|  | missense pLOF FAF | 1463 (7.8%) | 1374 (93.92%) | 84 (5.74%) | NA | NA | 5 (0.34%) |
| **Subclinical phenotype (ECG + CMR)** | Total | 18665 | 15056 (80.66%) | 2253 (12.07%) | 522 (2.8%) | 645 (3.46%) | 189 (1.01%) |
|  | missense pLOF FAF | 1463 (7.8%) | 1147 (78.4%) | 194 (13.26%) | 35 (2.39%) | 63 (4.31%) | 24 (1.64%) |
| **Combined phenotype** | Total | 18665 | 14578 (78.1%) | 2725 (14.6%) | 522 (2.8%) | 635 (3.4%) | 205 (1.1%) |
|  | missense pLOF FAF | 1463 (7.8%) | 1117 (76.35%) | 223 (15.24%) | 35 (2.39%) | 59 (4.03%) | 29 (1.98%) |

The number and proportion for each phenotype is shown for those with a pathogenic or likely pathogenic variant by the missense pLOF FAF filtering strategy and in the total study population for comparison.

**Table 3.** Demographic and clinical characteristics of the overall study population, putative pathogenic variant carriers (G+), and those without any putative pathogenic variants in DCM-associated genes (G-) for the primary variant filtering strategy.

|  | missense pLOF FAF |  |  |  |
| --- | --- | --- | --- | --- |
|  | Overall | G- | G+ | P |
| N | 18665 | 17202 | 1463 |  |
| Female, n (%) | 9844 (52.7) | 9076 (52.8) | 768 (52.5) | 0.866 |
| Age at recruitment, yrs | 54.96 (7.50) | 54.94 (7.49) | 55.16 (7.56) | 0.283 |
| Age at last follow up, yrs | 64.38 (7.47) | 64.37 (7.46) | 64.53 (7.58) | 0.436 |
| Systolic BP automated reading (mmHg), mean (SD) | 136.75 (18.66) | 136.75 (18.67) | 136.67 (18.48) | 0.866 |
| Diastolic BP automated reading (mmHg), mean (SD) | 81.25 (10.37) | 81.27 (10.39) | 81.09 (10.14) | 0.544 |
| Body mass index (kg/m<sup>2</sup>) | 26.51 (4.16) | 26.51 (4.17) | 26.53 (4.07) | 0.873 |
| Body surface area (m<sup>2</sup>) | 1.86 (0.21) | 1.86 (0.21) | 1.86 (0.21) | 0.865 |
| Creatinine (μmol/L) | 72.12 (14.09) | 72.11 (14.06) | 72.31 (14.45) | 0.601 |
| Dead, n (%) | 221 (1.2) | 204 (1.2) | 17 (1.2) | 1 |
| Age at death, yrs | 70.73 (6.71) | 70.64 (6.77) | 71.80 (5.98) | 0.493 |
| Coronary artery disease, n (%) | 846 (4.5) | 778 (4.5) | 68 (4.6) | 0.876 |
| Heart failure, n (%) | 240 (1.3) | 210 (1.2) | 30 (2.1) | 0.01 |
| Early DCM features, n (%) |  |  |  | 0.14 |
| Arrhythmia and/or CCD | 2729 (14.6) | 2505 (14.6) | 224 (15.3) |  |
| Isolated ventricular dilatation | 522 (2.8) | 487 (2.8) | 35 (2.4) |  |
| Hypokinetic non-dilated cardiomyopathy | 645 (3.5) | 582 (3.4) | 63 (4.3) |  |
| DCM overall, n (%) | 205 (1.1) | 176 (1.0) | 29 (2.0) | 0.00073 |
| Subclinical DCM | 189 (1.0) | 165 (1.0) | 24 (1.6) |  |
| Clinical DCM | 19 (0.1) | 14 (0.1) | 5 (0.3) |  |
| LVESV (ml) | 63.46 (63.75) | 63.50 (65.77) | 62.92 (31.60) | 0.737 |
| LVEDV (ml) | 140.72 (136.83) | 140.91 (141.97) | 138.57 (43.22) | 0.53 |
| LVEF (%) | 55.67 (6.62) | 55.69 (6.60) | 55.39 (6.81) | 0.094 |
| Hypertension, n (%) | 6028 (32.3) | 5574 (32.4) | 454 (31.0) | 0.295 |
| Stroke, n (%) | 330 (1.8) | 301 (1.7) | 29 (2.0) | 0.586 |
| Valvular heart disease, n (%) | 527 (2.8) | 487 (2.8) | 40 (2.7) | 0.894 |
| Atrial fibrillation/flutter, n (%) | 832 (4.5) | 765 (4.4) | 67 (4.6) | 0.865 |
| Bradyarrhythmia, n (%) | 352 (1.9) | 323 (1.9) | 29 (2.0) | 0.856 |
| Cardiac conduction defect, n (%) | 2448 (13.1) | 2238 (13.0) | 210 (14.4) | 0.155 |
| Sick sinus syndrome, n (%) | 19 (0.1) | 17 (0.1) | 2 (0.1) | 0.993 |
| CIED, n (%) | 148 (0.8) | 133 (0.8) | 15 (1.0) | 0.373 |
| Pre-excitation syndrome, n (%) | 35 (0.2) | 32 (0.2) | 3 (0.2) | 1 |
| Ventricular arrhythmia, n (%) | 69 (0.4) | 62 (0.4) | 7 (0.5) | 0.624 |
| eGFR (ml/min/1.73m<sup>2</sup>) | 87.32 (16.24) | 87.34 (16.36) | 87.10 (14.82) | 0.609 |
| CKD ≥3, n (%) | 396 (2.2) | 365 (2.2) | 31 (2.2) | 1 |

BP = blood pressure; DCM = dilated cardiomyopathy; CCD = cardiac conduction disease; CIED = cardiac implantable electronic device (including single- and dual-chamber permanent pacemaker, implantable cardioverter defibrillator, cardiac resynchronization therapy); CKD = chronic kidney disease; eGFR = estimated glomerular filtration rate; LVEDV = left ventricular end diastolic volume; LVESV = left ventricular end systolic volume; LVEF = left ventricular ejection fraction; SD = standard deviation.

### 3.5. Subclinical DCM and early DCM features in individuals with putative pathogenic variants

When assessed based on the CMR data, 24 (1.6%) additional individuals with putative pathogenic variants met the diagnostic criteria for DCM (subclinical DCM). There were no differences in the frequency of early DCM features between genotype-positive and genotype-negative groups.

### 3.6. Combined clinical and subclinical penetrance for DCM and early DCM-associated features

Overall, 346 (23.7%) individuals with putative pathogenic variants had DCM or showed early phenotypic features that may in part be attributed to DCM, most frequently arrhythmia and/or cardiac conduction disease (n=223, 64%). The overall penetrance of putative pathogenic variants combined for subclinical/clinical DCM and early DCM features, varied between 0% and 66.7% in those with single putative pathogenic gene variants and between 0% and 100% in those with two or more putative pathogenic variants. Overall, individuals with putative pathogenic variants more frequently developed a clinical or subclinical DCM phenotype, as compared with those without putative pathogenic variants (2% vs 1%, p=0.00073, Table 3).

### 3.7. Penetrance analysis based on gene-evidence category

A gene-evidence cluster-based analysis revealed slightly higher frequency of clinical DCM in participants with putative pathogenic variants in definitive/strong evidence genes, as compared to those with variants in moderate/limited evidence genes. However, combined clinical and subclinical phenotype was not statistically different between groups stratified based on gene-evidence category (Table 4).

**Table 4.** Clinical and subclinical phenotype in participants with putative pathogenic variants in DCM genes according to the pLOF missense FAF classification, stratified based on gene-evidence class.^1^.

|  | **Definitive/**<br>**Strong** | **Limited** | **Moderate** | **p-value\*** | **p-value\*\*** |
| --- | --- | --- | --- | --- | --- |
| N of participants, n (%) | 608 | 770 | 85 |  |  |
| Clinical phenotype, n (%) |  |  |  | 0.133 | 0.029 |
| Phenotype negative | 568 (93.4) | 726 (94.3) | 80 (94.1) |  |  |
| Arrhythmia and/or CCD | 35 (5.8) | 44 (5.7) | 5 (5.9) |  |  |
| Dilated cardiomyopathy | 5 (0.8) | 0 (0.0) | 0 (0.0) |  |  |
| Subclinical phenotype (ECG + CMR), n (%) |  |  |  | 0.542 | 0.488 |
| Phenotype negative | 480 (78.9) | 601 (78.1) | 66 (77.6) |  |  |
| Arrhythmia and/or CCD | 74 (12.2) | 109 (14.2) | 11 (12.9) |  |  |
| Isolated ventricular dilatation | 12 (2.0) | 22 (2.9) | 1 (1.2) |  |  |
| Hypokinetic non-dilated cardiomyopathy | 31 (5.1) | 28 (3.6) | 4 (4.7) |  |  |
| Dilated cardiomyopathy | 11 (1.8) | 10 (1.3) | 3 (3.5) |  |  |
| Combined phenotype, n (%) |  |  |  | 0.449 | 0.337 |
| Phenotype negative | 468 (77.0) | 584 (75.8) | 65 (76.5) |  |  |
| Arrhythmia | 85 (14.0) | 126 (16.4) | 12 (14.1) |  |  |
| Isolated ventricular dilatation | 12 (2.0) | 22 (2.9) | 1 (1.2) |  |  |
| Hypokinetic non-dilated cardiomyopathy | 27 (4.4) | 28 (3.6) | 4 (4.7) |  |  |
| Dilated cardiomyopathy | 16 (2.6) | 10 (1.3) | 3 (3.5) |  |  |

CCD = cardiac conduction disease.
<sup>1</sup> For the purpose of analysis, patients with >1 putative pathogenic variants in genes classified in different evidence categories were considered in the higher evidence category.

### 3.8. Prevalence of DCM-associated putative pathogenic gene variants using primary and secondary variant calling strategies

One or more putative pathogenic variant(s) in DCM-associated genes were identified in 1463 (7.8%) with the “missense pLOF FAF” variant calling strategy, in 154 (0.8%) using the “InterVar FAF” strategy and in 212 (1.1%) using the “InterVar FAF ClinVar” strategy (**Table S4**). The rate of diagnosis of clinical DCM varied between 0.3 and 1.4%. Early clinical features DCM (5.7-7.6%) were present in <8% of individuals carrying putative pathogenic variants; an additional 16.89-17.6% had a subclinical phenotype on ECG and/or CMR using different variant calling strategies. With a combined clinical/subclinical prevalence of 12.3-15.6%, arrhythmias and/or conduction disease were the most common early DCM features, followed by hypokinetic non-dilated cardiomyopathy, observed in 4.0-7.8%. The combined clinical/subclinical penetrance was ≤30% for all variant filtering strategies (Table 2 and **Table S4**).

### 3.9. Gene-based analysis of penetrance and clinical phenotype

Overall, putative pathogenic variants were found in all screened 44 genes by the missense pLOF FAF strategy, and in a lower number of genes when using the more strict secondary strategies. The gene-specific penetrance ranged from 0 to 66.7% (Figure 2). A subanalysis for genes *TNNT2, LMNA, FLNC*, and *TTN*, which are listed in the ACMG clinically actionable gene list, revealed DCM or early DCM features in respectively 10.5%, 26.7%, 22.3%, and 45.4% of cases, indicating higher penetrance than in the overall putative pathogenic population. Gene-based analysis of penetrance for additional filtering strategies is provided in **Figure S2** and **Tables S5-S7**.

### 3.10. Impact of DCM-associated putative pathogenic variants on clinical outcomes

In order to assess the impact of DCM-associated putative pathogenic variants on the clinical outcome, an event-free survival analysis was performed for genotype-positive versus genotype negative patients for each of the three variant filtering strategies. Event-free survival was defined as survival without developing one of heart failure, stroke, arrhythmia, required CIED, or death. There was no statistical difference in the survival between individuals with and without putative pathogenic variants (HR 1.06 [95%CI 0.87 – 1.29] for the primary strategy; **Figure S3**). Additional analysis regarding prevalence and incidence of DCM and potentially DCM-associated clinical features in genotype-positive and genotype-negative groups are provided in **Tables S8-S9**.

## 4. DISCUSSION

Using the UK Biobank WES data, we analyzed the prevalence and penetrance of DCM-associated gene variants in a cohort of over 18,000 individuals with 12-lead ECG and CMR. This is the first study providing insights into the clinical and subclinical penetrance of DCM-associated gene variants in a large population-scale dataset and has several important findings with direct clinical implications. First, the UK Biobank population of mainly middle-aged adults has a prevalence of non-ischemic DCM of 1:355 (0.28%). Second, the variant filtering strategy used for those with a clinical diagnosis of DCM provided a yield of 16%. Third, using the same strategy and a genotype-first approach identified 1,463 (7.8%) individuals with putative pathogenic DCM gene variants. Fourth, clinical/subclinical disease penetrance was highly variable, ranging from 0 to 66.7% between genes. Fifth, among individuals with putative pathogenic DCM variants, subclinical DCM and early DCM features, detected by 12-ECG and/or CMR, were five times more common than clinically manifest DCM (21.6% vs. 3.8%; p<0.00001) (Figure 3). Finally, participants with putative pathogenic variants in definitive/strong DCM genes appeared to have slightly higher rate of clinical DCM, than those with variants in lower evidence genes, but combined clinical and subclinical phenotype was not statistically different between groups stratified based on gene-evidence categories.

**Figure 3.**
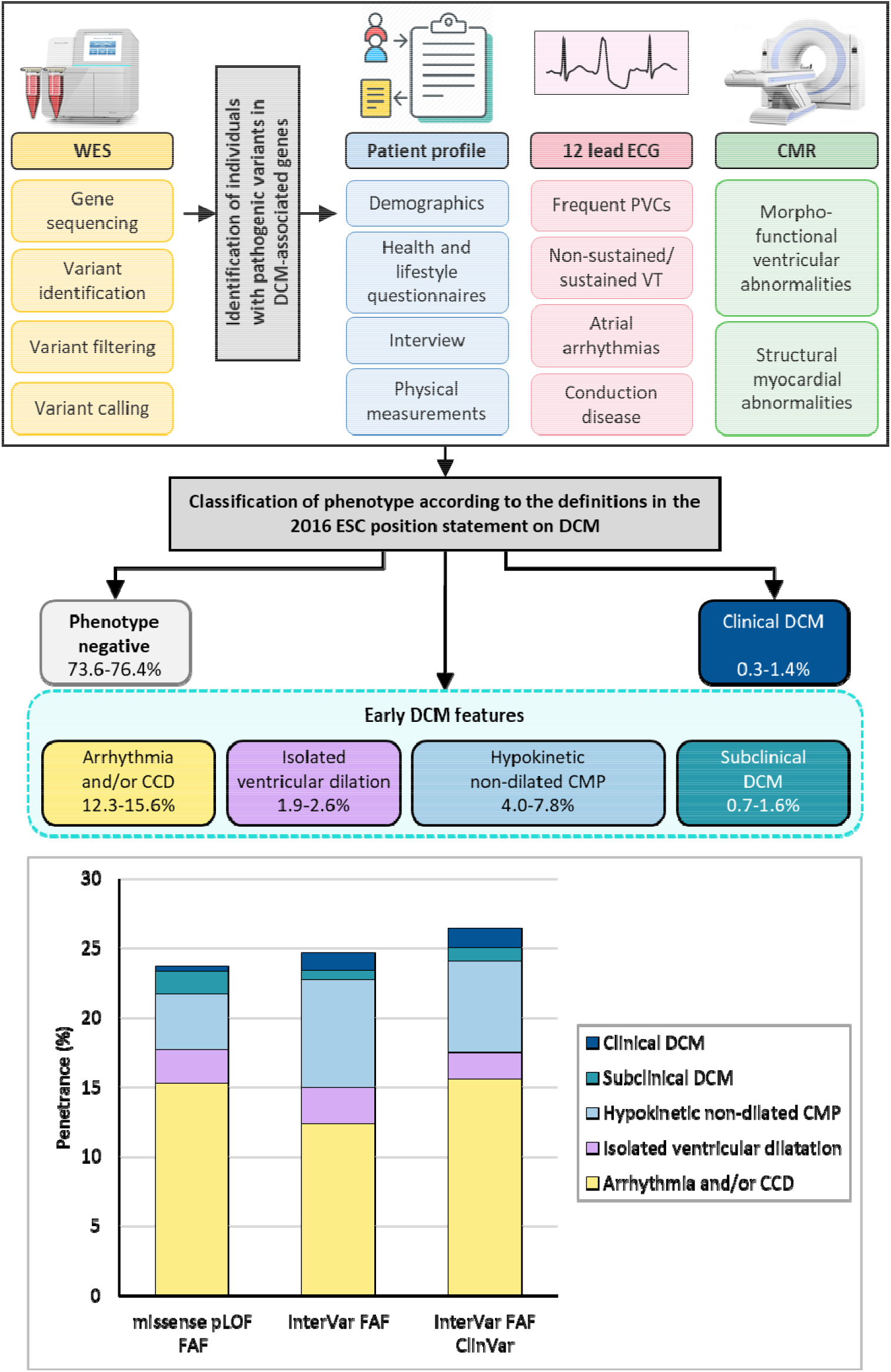
Central illustration of the study, demonstrating the methodology used for phenotype ascertainment, classification and clinical/subclinical phenotype in participants with putative pathogenic variants in DCM genes according to the three variant filtering strategies applied.

Our analysis indicates a DCM prevalence of 1:355 in the UK Biobank population, with over 3.3:1 male predominance, in line with previous population-based epidemiological studies of non-ischemic DCM.33, 34 Studies reporting a prevalence of 1 in 250 have, perhaps, included all etiologies of a morphological DCM phenotype. Among individuals with putative pathogenic variants in the screened DCM-associated genes, only 0.3% had a clinically diagnosed DCM, but an additional 1.6% (5-fold increase) were found to meet the CMR criteria for DCM, indicating that most DCM cases in the general adult population go unnoticed for many years. Interestingly, the 12-lead ECG and CMR screening in those with putative pathogenic variants identified 16.2% of individuals with subclinical early features of DCM, such as cardiac arrhythmias, isolated ventricular dilation, and hypokinetic non-dilated DCM. In fact, the population with early DCM features was 11-fold larger than those with clinical and subclinical DCM combined (21.7% vs. 2%), demonstrating the wide variability of phenotypic expression in individuals with putative pathogenic variants in DCM genes. Arrhythmias and CCD were the most common early manifestation of putative pathogenic variants implicated in DCM, mostly occurring prior to the development of structural/functional abnormalities. These findings are in line with observations in the large and well-phenotyped Geisinger database, showing very low penetrance of arrhythmogenic right ventricular cardiomyopathy-associated *PKP2* variants in the middle-aged population.35

An unsolved challenge in clinical genetics is that the principles applied to variant calling are probabilistic. While the 2015 ACMG guidelines integrate a robust data scheme to classify gene variants,36 applying these criteria to biobank studies is challenging because expert-curator input is often necessary to assess variant pathogenicity and gene-disease association. Here, we applied three special variant calling models developed for this study to all genes, except for *TTN*, where we only considered radical variants as putative pathogenic variants. The two secondary filtering strategies we used can be considered more stringent than the primary strategy; these yielded a lower frequency of variants in the studied population with a similar low penetrance, indicating that the true DCM penetrance is very low regardless of variant calling strategy. On the other hand, many genes, such as *SCN5A, DSP, MYH7, DES*, and others, show broad pleiotropy and variability of phenotypic expression, making the definition of positive phenotype in large datasets rather complicated. In order to extend the breadth of recognizable phenotypes we included early features of DCM. However, it is possible that the individuals with putative pathogenic variants in DCM genes, who showed no DCM features, manifest other cardiomyopathy phenotypes, or in the case of *SCN5A*, primary arrhythmia syndromes.37

Of the genes included in the ClinGen DCM panel used in this study to define a DCM-relevant gene panel, *TNNT2, LMNA, FLNC*, and *TTN* are among the 59 genes identified as medically actionable by the ACMG,13 for which clinical management guidelines have been established. In the UK Biobank subcohort, individuals with putative pathogenic variants in the *TNNT2, LMNA, FLNC* and *TTN* genes showed signs of DCM or early DCM features in respectively 10.5%, 26.7%, 22.3%, and 45.4% of cases. Indeed, this is significantly higher penetrance than that seen in the overall DCM gene panel.

Although individuals with putative pathogenic variants in DCM genes had a markedly higher observed frequency of clinical/subclinical DCM and two-fold higher frequency of heart failure, our analysis reflects low penetrance of DCM-associated putative pathogenic variants in a middle to older aged individuals, indicating that most of these individuals are unlikely to develop disease. The presence of a putative pathogenic variant in the DCM-associated genes was not associated with a higher risk of heart failure or worse outcome, similar to the findings of Carruth et al. for pathogenic alleles in the *PKP2* gene.35 It should be noted that monogenic disease expressivity and penetrance, particularly in DCM, is dependent on genomic context, as indicated by family history or the effect of polygenic risk,38, 39 as well as the environment (e.g. alcohol and pregnancy in *TTNtv*-mediated DCM, inflammation in *DSP-*mediated cardiomyopathy).40-43 Future studies investigating population penetrance of DCM should consider disease modifiers and approach to DCM as a multifactorial trait highly influenced by environmental and genetic modifiers.

A population-based ‘genotype first’ screening strategy must fulfill certain criteria in order to be cost-effective.44 First, the condition should be a sufficient health problem, which DCM is. Second, the natural history should be understood. In DCM, this is in part understood, but as multimodality imaging and physician awareness improve, we have learned that the natural history can be quite different for this genetically heterogeneous group. Third, there should be a recognizable latent or early symptomatic stage, which for heart failure, isolated ventricular dilation and hypokinetic non-dilated cardiomyopathy is applicable to DCM. However, for those in whom the sentinel event is sudden death, this is not fulfilled. Fourth, there should be a suitable test or examination, which is fulfilled by echocardiography and CMR. Fifth, screening should be acceptable to the population, and, in general, cardiovascular diseases are. Sixth and seventh, there should be an agreed policy on whom to treat and expected treatment, both of which are well satisfied regarding established heart failure syndrome and arrhythmia, but not agreed on asymptomatic mild or subclinical disease. Eighth, facilities for diagnosis and treatment should be available, which has been achieved throughout most of the developed world, though disparities persist and remain lacking in the developing nations.45 Ninth, the cost of case finding, diagnosis, and treatment, should be economically balanced, which with clinical diagnosis is justified, but for genetics, it is difficult to justify given upfront costs and the low signal-to-noise ratio. This may be improved with a targeted panel of very high-risk genes and higher penetrance. In our study, only *TTN*tv would meet this criteria, whereas genes considered as high-risk in consensus statements and guidelines, such as *DSP, LMNA, RBM20, SCN5A, FLNC*, and *PLN*,46 had a low frequency of putative pathogenic variants and low penetrance. Based on current data, a genotype-first screening strategy in DCM would be difficult to justify due to high cost and low penetrance. However, this should be interpreted in the context of this dataset being a relatively healthy and older population, where high-risk individuals may have succumbed to sudden death or not enrolled. Lastly, case finding should be a continuous process, and applying this to clinical screening for presentations of DCM may be better achieved through phenotype screening tools such as artificial intelligence–enabled ECG.47 Years later, this methodology may be a more cost-effective approach than a genetic screening tool and then continuous clinical phenotyping, which are currently prohibitively expensive.

### Strengths and Limitations

Our study has a number of strengths. First, we used a robust methodology with one primary and two secondary variant calling strategies, which all confirmed the low yield of clinical and subclinical DCM in putative pathogenic variant carriers. Second, in order to include early DCM features as defined by Pinto et al.23, as well as those with subclinical DCM, we analyzed electronic health records, 12-lead ECG and CMR data to enable the deeper phenotyping of individuals. Of note, this is the first study to report detailed high throughput computer interpretation, with manual validation, of 12-lead ECGs from the UK Biobank population. Third, our study included a cohort of over 18,000 individuals who underwent CMR according to a standardized protocol.

The study results should be viewed in light of several limitations. First, some DCM genes show significant pleiotropy with the development of distinct phenotypes. We did not consider phenotypic features other than DCM or early DCM features. This may have resulted in underestimation of the clinical penetrance of these genes when it relates to other phenotypes. Second, the UK Biobank population may reflect volunteer bias and survivor bias with a sample of healthier individuals than the general UK population,48 and thus may show lower frequency of putative pathogenic variants and lower penetrance. Third, the ethnicity is mainly White British individuals, making generalizability to other ethnicities challenging, particularly in a disease with known differences in genetic etiology in different races.49, 50 Fourth, although 200,000 WES data are available, only 18,000 had both 12-lead ECG and CMR available (which were usable); with the staged approach, and impacts of the pandemic, the goals of 500,000 WES and 100,000 with 12-lead ECG and CMR are unlikely to be achieved on time. A larger sample of 100,000 and with longer follow-up may show higher penetrance, given the age of onset of DCM is also variable. The UK Biobank does not include recruits aged less than 40 years, which may underestimate prevalence, and exclude high-risk DCM patients who succumb to sudden death events. Detection of late gadolinium enhancement is not part of UK Biobank CMR protocol; right ventricular phenotypic expression was not evaluated for this study.

## 5. CONCLUSIONS

Over 90% of middle and older aged adults with putative pathogenic variants in DCM-associated genes did not have a history of DCM or of early DCM features. Nearly one-sixth of putative pathogenic variant carriers exhibited a subclinical phenotype on ECG and/or CMR, most commonly manifesting with arrhythmias in the absence of substantial ventricular dysfunction. Given the difficulties in variant pathogenicity adjudication, low disease penetrance and uncertainties in clinical actionability, applying a gene-first approach to DCM for clinical and investigative decision making might currently be challenging for a broad gene panel, but might be useful for clinically actionable genes, which show a relatively higher penetrance.

## Data Availability

All data produced in the present work are contained in the manuscript.

## Acknowledgments

This study was conducted using the UK Biobank resource under access application 48286.

We would like to thank all the participants, staff involved with planning, collection and analysis, including core lab analysis of the CMR imaging data.

## Sources of Funding

APL is supported by the American Academy of Pediatrics, Children’s Cardiomyopathy Foundation, and the Derfner Foundation. AML reports funding from UKRI London Medical Imaging & Artificial Intelligence Centre for Value Based Healthcare and SmartHeart” EPSRC program grant (EP/P001009/1). LRL is funded by MRC UK Clinical Academic Research Partnership award (MR/T005181/1). CS is the recipient of a National Health and Medical Research Council (NHMRC) Practitioner Fellowship (#1154992). SEP acknowledge the British Heart Foundation for funding the manual analysis to create a cardiovascular magnetic resonance imaging reference standard for the UK Biobank imaging resource in 5000 CMR scans (www.bhf.org.uk; PG/14/89/31194). NA recognises the National Institute for Health Research (NIHR) Integrated Academic Training programme, which supports his Academic Clinical Lectureship post. SEP and PBM acknowledge support from the National Institute for Health Research (NIHR) Biomedical Research Centre at Barts. SEP acknowledges support from and from the “SmartHeart” EPSRC programme grant (www.nihr.ac.uk; EP/P001009/1). This project was enabled through access to the MRC eMedLab Medical Bioinformatics infrastructure, supported by the Medical Research Council (www.mrc.ac.uk; MR/L016311/1). The funders provided support in the form of salaries for authors as detailed above, but did not have any additional role in the study design, data collection and analysis, decision to publish, or preparation of the manuscript.

## Disclosures

Dr. Lopes serves in the advisory board of BMS. Dr. Owens provided consulting to BMS and Cytokinetics. Dr. Petersen provided consultancy to and is shareholder of Circle Cardiovascular Imaging Inc., Calgary, Alberta, Canada. The other authors report that they have no relevant conflicts of interest to disclose.

## Notes

### Funding Statement

APL is supported by the American Academy of Pediatrics, Childrens Cardiomyopathy Foundation, and the Derfner Foundation. AML reports funding from UKRI London Medical Imaging & Artificial Intelligence Centre for Value Based Healthcare and SmartHeart EPSRC program grant (EP/P001009/1). LRL is funded by MRC UK Clinical Academic Research Partnership award (MR/T005181/1). CS is the recipient of a National Health and Medical Research Council (NHMRC) Practitioner Fellowship (#1154992). SEP acknowledge the British Heart Foundation for funding the manual analysis to create a cardiovascular magnetic resonance imaging reference standard for the UK Biobank imaging resource in 5000 CMR scans (www.bhf.org.uk; PG/14/89/31194). NA recognises the National Institute for Health Research (NIHR) Integrated Academic Training programme, which supports his Academic Clinical Lectureship post. SEP and PBM acknowledge support from the National Institute for Health Research (NIHR) Biomedical Research Centre at Barts. SEP acknowledges support from and from the SmartHeart EPSRC programme grant (www.nihr.ac.uk; EP/P001009/1). This project was enabled through access to the MRC eMedLab Medical Bioinformatics infrastructure, supported by the Medical Research Council (www.mrc.ac.uk; MR/L016311/1). The funders provided support in the form of salaries for authors as detailed above, but did not have any additional role in the study design, data collection and analysis, decision to publish, or preparation of the manuscript.

### Author Declarations

This study involves only openly available human data, which can be obtained from https://www.ukbiobank.ac.uk/

