## Supplementary Material for "The Frequency, Penetrance and Variable Expressivity of Dilated Cardiomyopathy-Associated Putative Pathogenic Gene Variants in UK Biobank Participants"

**Supplemental Methods**

**Variant Filtering**

In addition to the main filtering strategy: “missense pLOF FAF”, two other variant filtering strategies were performed. The first of these (“InterVar FAF”) used InterVar, a tool that generates automated variant interpretation using the 2015 American College of Medical Genetics and Genomics (ACMG) and the Association for Molecular Pathology criteria.1 Variants with InterVar classifications of pathogenic or likely pathogenic and a filtering allele frequency (FAF) ≤ $8.4 \times{10}^{-5}$ formed the set of putative pathogenic variants.

In the final filtering strategy (“InterVar FAF ClinVar”), variants were categorized as putative pathogenic if they had been identified in the second filtering strategy (“InterVar FAF”) and/or listed in ClinVar as pathogenic or likely pathogenic with at least 2-star evidence level.

All three filtering strategies were also applied to cases with a clinical diagnosis of dilated cardiomyopathy (DCM) in UK Biobank, where whole exome sequencing was available, to determine yield. Therefore, this provided a quality control measure of the filtering strategies.

**Cardiac Magnetic Resonance (CMR) Analysis**

Manual analysis of left ventricular volumes (LV) and mass were performed as per international recommendations.2 As part of the UK Biobank CMR analysis, D13A InlineVF analysis algorithm is provided, with separate DICOM image storage.3 This was updated with a subsequent software release E11c which provides LV mass based on epicardial contouring. In brief, this uses the shortest algorithms to determine endocardial and epicardial contours propogated to other frames and in other slices; long axis images are used to identify basal and apical landmarks using machine learning methods. Papillary muscles were excluded from LV mass calculations and considered as part of the blood pool. Outliers with end diastolic or end systolic volumes >500 ml were excluded from automated analyses. Eight readers in two core laboratories were trained using standard operating procedures to ensure minimal inter- and intra-observer bias. CMR examinations were analyzed using Circle CVI post-processing software (Version 5.1.1, Circle Cardiovascular Imaging Inc., Calgary, Canada), excluding papillary muscles. The end diastolic frame was selected as the first frame of the series; and the end systolic frame selected as the smallest LV blood pool area in the mid-ventricular slice. Software provided determinants of LV mass and volumes, assuming myocardial density of 1.06 g/ml for both the InlineVF method and Circle CVI. Analysis showed that fully automated InlineVF algorithm performed as well as semi-automated.

**Statistical Analysis**

To test for significant differences in outcomes between genotype positive and genotype negative individuals Cox proportional hazards regression was used. Survival without developing one of heart failure, stroke, arrhythmia, required cardiac implantable electronic device (including single- and dual-chamber permanent pacemaker, implantable cardioverter defibrillator, cardiac resynchronization therapy), or death was used as the outcome. Survival duration was calculated from date of enrollment to date of censoring. Data was treated as left truncated (by date of enrollment) and right censored. Age was chosen as the timescale rather than time-on-study as this approach reduces bias associated with potential confounding by age in cohort studies4^,^ 5. This analysis was performed using the *survival* package.

**Table S1. Table of phenotypic definitions used.**

| **Phenotype** | **Data fields** | **Field names** | **Data codes** | **Data code definitions** |
| --- | --- | --- | --- | --- |
| Atrial fibrillation or flutter | 20002 | Non-cancer illness code, self-reported | 1471, 1483 | Atrial fibrillation, Atrial flutter |
| Atrial fibrillation or flutter | 20004 | Operation code, self-reported | 1524 | Cardioversion |
| Atrial fibrillation or flutter | 41202  41204  40001  40002 | Diagnoses - main ICD10  Diagnoses – secondary ICD10  Underlying (primary) cause of death: ICD10  Contributory (secondary) cause of death: ICD10 | I48, I48.0, I48.1, I48.2, I48.3, I48.4, I48.9 | Atrial fibrillation and flutter, Paroxysmal atrial fibrillation, Persistent atrial fibrillation, Chronic atrial fibrillation, Typical atrial flutter, Atypical atrial flutter, Atrial fibrillation and atrial flutter, unspecified |
| Atrial fibrillation or flutter | 41203  41205 | Diagnoses - main ICD9  Diagnoses - secondary ICD9 | 4273 | Atrial fibrillation and flutter |
| Atrial fibrillation or flutter | 41200  41210 | Operative procedures - main OPCS Operative procedures - secondary OPCS | K57.1, K62.1, K62.2, K62.3, K62.4 | Percutaneous transluminal ablation of atrioventricular node, Percutaneous transluminal ablation of pulmonary vein to left atrium conducting system, Percutaneous transluminal ablation of atrial wall for atrial flutter,  Percutaneous transluminal ablation of conducting system of heart for atrial flutter NEC, Percutaneous transluminal internal cardioversion NEC |
| Atrial fibrillation or flutter | ECG analysis | MUSE definitions | 161, 162, 273, 288 | Atrial fibrillation, Atrial flutter, Atrial flutter, Atrial flutter with 2 to 1 block |
| Bradyarrhythmias | 20002 | Non-cancer illness code, self-reported | 1486 | Sick sinus syndrome |
| Bradyarrhythmias | 41202  41204  40001  40002 | Diagnoses - main ICD10  Diagnoses – secondary ICD10  Underlying (primary) cause of death: ICD10  Contributory (secondary) cause of death: ICD10 | I44, I44.1, I44.2, I44.3, I44.5, I49.5 | Atrioventricular and left bundle-branch block, Atrioventricular block, second degree, Atrioventricular block, complete, Other and unspecified atrioventricular block, Other specified heart block, Sick sinus syndrome |
| Bradyarrhythmias | 41203  41205 | Diagnoses - main ICD9  Diagnoses - secondary ICD9 | 4260, 4261, 4266 | Atrioventricular block, complete, Atrioventricular block, other and unspecified, Other specified heart block |
| Bradyarrhythmias | 20004 | Operation code, self-reported | 1096, 1548, 1549 | Pacemaker/defibrillator insertion, Pacemaker insertion, Pacemaker battery change |
| Bradyarrhythmias | 3079 | Pace-maker, verbal interview | 1 | Yes |
| Bradyarrhythmias | 41200  41210 | Operative procedures - main OPCS Operative procedures - secondary OPCS | K60, K60.1, K60.2, K60.3, K60.4, K60.5, K60.6, K60.8, K60.9, K61, K61.1, K61.2,  K61.3, K61.4, K61.5, K61.6, K61.8, K61.9 | Cardiac pacemaker system introduced through vein, Implantation of intravenous cardiac pacemaker system NEC, Resiting of lead of intravenous cardiac pacemaker system, Renewal of intravenous cardiac pacemaker system, Removal of intravenous cardiac pacemaker system, Implantation of intravenous single chamber cardiac pacemaker system, Implantation of intravenous dual chamber cardiac pacemaker system, Other specified cardiac pacemaker system introduced through vein, Unspecified cardiac pacemaker system introduced through vein, Other cardiac pacemaker system, Implantation of cardiac pacemaker system NEC, Resiting of lead of cardiac pacemaker system NEC, Renewal of cardiac pacemaker system  NEC, Removal of cardiac pacemaker system NEC, Implantation of single chamber cardiac pacemaker system, Implantation of dual chamber cardiac  pacemaker system, Other specified other cardiac pacemaker system, Unspecified other cardiac pacemaker system |
| Bradyarrhythmia | ECG analysis | MUSE definitions | 104, 106, 107, 113, 141, 142, 143, 144, 281, 285, | with 2nd degree AV block (Mobitz II), with complete heart block, with variable AV block, with sinus pause, with 2:1 AV conduction, with 3:1 AV conduction, with 4:1 AV conduction, with 5:1 AV conduction, Complete heart block, with sinus arrest or transient AV block |
| Conduction system diseases | 41202  41204  40001  40002 | Diagnoses - main ICD10  Diagnoses – secondary ICD10  Underlying (primary) cause of death: ICD10  Contributory (secondary) cause of death: ICD10 | I44.0, I44.4, I44.5, I44.6, I44.7, I45.0, I45.1, I45.2, I45.3, I45.4 | Atrioventricular block, first degree, Left anterior fascicular block, Left posterior fascicular block, Other and unspecified fascicular block, Left bundle-branch block, unspecified, Right fascicular block, Other and unspecified right bundle- branch block, Bifascicular block, Trifascicular block, Nonspecific intraventricular block |
| Conduction system diseases | 41203  41205 | Diagnoses - main ICD9  Diagnoses - secondary ICD9 | 4263, 4264, 4265 | Other left bundle branch block, Right bundle branch block, Bundle branch block, unspecified |
| Conduction system diseases | ECG analysis | MUSE definitions | 101, 470, 371, 372, 471, 460, 440, 478, 479, 480, 481, 482, 487 | with 1st degree AV block, Left anterior fascicular block, abnormal left axis deviation, left axis deviation, left posterior fascicular block, left bundle branch block, right bundle branch block, RBBB and left anterior fascicular block, RBBB and left posterior fascicular block, Bifascicular block, Trifascicular block, Nonspecific intraventricular block, Nonspecific intraventricular conduction delay, QRS>110ms |
| Coronary artery disease | 41202  41204  40001  40002 | Diagnoses - main ICD10  Diagnoses – secondary ICD10  Underlying (primary) cause of death: ICD10  Contributory (secondary) cause of death: ICD10 | I21.X, I22.X, I23.X, I24.1, I25.2 | Myocardial infarction |
| Coronary artery disease | 20002 | Non-cancer illness code, self-reported | 1075 | Heart attack/myocardial infarction |
| Coronary artery disease | 41200  41210 | Operative procedures - main OPCS Operative procedures - secondary OPCS | K40.1–40.4, K41.1–41.4, K45.1–45.5, K49.1–49.2, K49.8–49.9, K50.2, K75.1–75.4, K75.8–75.9 | Coronary artery bypass grafting, coronary angioplasty with or without stenting |
| Coronary artery disease | 20004 | Operation code, self-reported | 1070, 1095 | coronary angioplasty (ptca) +/- stent, coronary artery bypass grafts (cabg) |
| Diabetes | 20002 | Non-cancer illness code, self-reported | 1220  1222  1223 | Diabetes  Type 1 diabetes  Type 2 diabetes |
| Diabetes | 41202  41204  40001  40002 | Diagnoses - main ICD10  Diagnoses – secondary ICD10  Underlying (primary) cause of death: ICD10  Contributory (secondary) cause of death: ICD10 | E10-14 | Diabetes mellitus |
| Dilated cardiomyopathy | 41202  41204  40001  40002 | Diagnoses - main ICD10 Diagnoses – secondary ICD10  Underlying (primary) cause of death:ICD10  Contributory (secondary) cause of death: ICD10 | I42.0 | Dilated cardiomyopathy |
| Heart failure | 20002 | Non-cancer illness code, self-reported | 1076, 1079 | Heart failure, cardiomyopathy |
| Heart failure | 41202  41204  40001  40002 | Diagnoses - main ICD10  Diagnoses – secondary ICD10  Underlying (primary) cause of death: ICD10  Contributory (secondary) cause of death: ICD10 | I11.0, I13.0, I13.2, I25.5, I42.0, I42.5, I42.8, I42.9, I50.0, I50.1, I50.9 | hypertensive heart disease, cardiomyopathy, heart failure |
| Hypertension | 20002 | Non-cancer illness code, self-reported | 1065, 1072 | Essential hypertension, hypertension |
| Hypertension | 41202  41204  40001  40002 | Diagnoses - main ICD10  Diagnoses – secondary ICD10  Underlying (primary) cause of death: ICD10  Contributory (secondary) cause of death: ICD10 | I10-5 | Hypertensive diseases |
| Supraventricular arrhythmias | 20002 | Non-cancer illness code, self-reported | 1484 | Wolff-Parkinson Wwhite / WPW syndrome |
| Supraventricular arrhythmias | 41202  41204  40001  40002 | Diagnoses - main ICD10  Diagnoses – secondary ICD10  Underlying (primary) cause of death: ICD10  Contributory (secondary) cause of death: ICD10 | I45.6 | Preexcitation syndrome |
| Supraventricular arrhythmias | 41203  41205 | Diagnoses - main ICD9  Diagnoses - secondary ICD9 | 4267 | Anomalous atrioventricular excitation |
| Supraventricular arrhythmias | 41200  41210 | Operative procedures - main OPCS  Operative procedures – secondary OPCS | K52.4,  K57.4 | Open division of accessory pathway within heart, Percutaneous transluminal ablation of accessory pathway, |
| Supraventricular arrhythmias | ECG analysis | MUSE definitions | 300, 302, 303, 304 | Ventricular pre-excitation, WPW pattern type A, Ventricular pre-excitation, WPW pattern type B, with fusion or intermittent ventricular pre-excitation (WPW), Wolff-Parkinson-White |
| Valvular heart disease | 41202  41204  40001  40002 | Diagnoses - main ICD10  Diagnoses – secondary ICD10  Underlying (primary) cause of death: ICD10  Contributory (secondary) cause of death: ICD10 | I34-37, I39.0-39.4, I05-08, I09.1, I09.8 | Aortic, mitral, pulmonary and tricuspid valve disorders |
| Valvular heart disease | 20002 | Non-cancer illness code, self-reported | 1078, 1488, 1584, 1585, 1489, 1586, 1587, 1490 | heart valve problem/heart murmur, mitral valve prolapse, mitral valve disease, mitral regurgitation/incompetence, mitral stenosis, aortic valve disease, aortic regurgitation/incompetence, aortic stenosis |
| Ventricular arrhythmias | 41202  41204  40001  40002 | Diagnoses - main ICD10  Diagnoses – secondary ICD10  Underlying (primary) cause of death: ICD10  Contributory (secondary) cause of death: ICD10 | I47.0, I47.2, I49.0, I46.0,  I46.1, I46.9 | Reentry ventricular arrhythmia, Ventricular tachycardia, Ventricular fibrillation and flutter, Cardiac arrest with successful resuscitation, Sudden cardiac  death, so described, Cardiac arrest, unspecified |
| Ventricular arrhythmias | 41203  41205 | Diagnoses - main ICD9  Diagnoses - secondary ICD9 | 4271, 4274, 4275 | Paroxysmal ventricular tachycardia, Ventricular fibrillation and flutter, Cardiac arrest |
| Ventricular arrhythmias | 41200  41210 | Operative procedures - main OPCS  Operative procedures – secondary OPCS | K57.6, K64.1, X50.3, X50.4, X50.8, X50.9 | Percutaneous transluminal ablation of ventricular wall NEC, Percutaneous radiofrequency ablation of epicardium, Advanced cardiac pulmonary resuscitation, External ventricular defibrillation, Other specified external resuscitation, Unspecified external resuscitation |

UK Biobank data field numbers and names are referenced along with the data codes and their associated definitions were appropriate.

Table adapted from eTable 2 and 3 in Khurshid et al. 20186, additional definitions from Khera et al. 2018 7 and Aragam 8 et al. 2018 where available.

### Table S2. Baseline characteristics of the overall UK Biobank population and of phenotypic subgroups, defined by clinical diagnoses.

|  | Overall | Phenotype negative | Clinical diagnosis of DCM | Clinical diagnosis of Arrhythmia and/or CCD |
| --- | --- | --- | --- | --- |
| N | 502462 | 451840 | 1415 | 49207 |
| Female, n (%) | 273354 (54.4) | 254697 (56.4) | 427 (30.2) | 18230 (37.0) |
| Age at recruitment, yrs | 56.53 (8.10) | 55.97 (8.07) | 59.80 (7.00) | 61.56 (6.41) |
| Age at last follow up, yrs | 65.68 (8.09) | 65.22 (8.06) | 68.77 (7.02) | 70.68 (6.57) |
| Systolic BP automated reading (mmHg) | 139.74 (19.70) | 139.20 (19.54) | 139.25 (21.28) | 144.66 (20.38) |
| Diastolic BP automated reading (mmHg) | 82.21 (10.70) | 82.16 (10.64) | 81.83 (11.93) | 82.66 (11.18) |
| Body mass index (kg/m^2^) | 27.43 (4.80) | 27.28 (4.72) | 29.41 (5.62) | 28.80 (5.28) |
| Body surface area (m^2^) | 1.86 (0.21) | 1.86 (0.21) | 2.02 (0.20) | 1.93 (0.20) |
| Creatinine (μmol/L) | 72.31 (18.55) | 71.65 (16.57) | 83.74 (34.74) | 78.03 (30.33) |
| Dead, n (%) | 35036 (7.0) | 24854 (5.5) | 381 (26.9) | 9801 (19.9) |
| Age at death, yrs | 69.73 (7.42) | 68.81 (7.59) | 69.62 (7.65) | 72.08 (6.38) |
| Coronary artery disease, n (%) | 36348 (7.2) | 22205 (4.9) | 482 (34.1) | 13661 (27.8) |
| Heart failure, n (%) | 16693 (3.3) | 5092 (1.1) | 1366 (96.5) | 10235 (20.8) |
| Hypertension, n (%) | 193754 (38.6) | 157924 (35.0) | 1094 (77.3) | 34736 (70.6) |
| Stroke, n (%) | 14777 (2.9) | 9611 (2.1) | 122 (8.6) | 5044 (10.3) |
| Valvular heart disease, n (%) | 20959 (4.2) | 8897 (2.0) | 593 (41.9) | 11469 (23.3) |
| Atrial fibrillation/flutter, n (%) | 35088 (7.0) | 0 (0.0) | 810 (57.2) | 34278 (69.7) |
| Bradyarrhythmia, n (%) | 17714 (3.5) | 0 (0.0) | 723 (51.1) | 16991 (34.5) |
| Cardiac conduction defect, n (%) | 13159 (2.6) | 0 (0.0) | 585 (41.3) | 12574 (25.6) |
| Sick sinus syndrome, n (%) | 1225 (0.2) | 0 (0.0) | 29 (2.0) | 1196 (2.4) |
| CIED, n (%) | 10337 (2.1) | 0 (0.0) | 412 (29.1) | 9925 (20.2) |
| Pre-excitation syndrome, n (%) | 532 (0.1) | 0 (0.0) | 11 (0.8) | 521 (1.1) |
| Ventricular arrhythmia, n (%) | 5090 (1.0) | 0 (0.0) | 281 (19.9) | 4809 (9.8) |

BP = blood pressure; DCM = dilated cardiomyopathy; CIED = cardiac implantable electronic device (including single- and dual-chamber permanent pacemaker, implantable cardioverter defibrillator, cardiac resynchronization therapy)

**Table S3. Demographic and clinical characteristics of the overall study population (with whole exome sequencing, usable cardiac magnetic resonance and 12-lead ECG), of putative pathogenic variant carriers (G+), and those without any putative pathogenic variants (G-) for each of the two secondary variant filtering strategies.**

|  |  | InterVar FAF | | | InterVar FAF ClinVar | | |
| --- | --- | --- | --- | --- | --- | --- | --- |
|  | Overall | G- | G+ | p | G- | G+ | p |
| **N** | 18665 | 18511 | 154 |  | 18453 | 212 |  |
| **Female, n (%)** | 9844 (52.7) | 9765 (52.8) | 79 (51.3) | 0.78 | 9740 (52.8) | 104 (49.1) | 0.312 |
| **Age at recruitment, yrs** | 54.96 (7.50) | 54.95 (7.50) | 55.66 (7.43) | 0.243 | 54.95 (7.50) | 55.51 (7.38) | 0.283 |
| **Age at last follow up, yrs** | 64.38 (7.47) | 64.38 (7.47) | 65.03 (7.54) | 0.283 | 64.38 (7.47) | 64.93 (7.43) | 0.279 |
| **Systolic BP automated reading (mmHg)** | 136.75 (18.66) | 136.74 (18.67) | 137.67 (17.32) | 0.545 | 136.74 (18.67) | 137.35 (17.66) | 0.643 |
| **Diastolic BP automated reading (mmHg)** | 81.25 (10.37) | 81.26 (10.37) | 80.73 (10.23) | 0.542 | 81.26 (10.38) | 81.12 (10.07) | 0.857 |
| **Body mass index (kg/m^2^)** | 26.51 (4.16) | 26.51 (4.16) | 26.61 (4.15) | 0.776 | 26.51 (4.16) | 26.49 (3.96) | 0.951 |
| **Body surface area (m^2^)** | 1.86 (0.21) | 1.86 (0.21) | 1.85 (0.21) | 0.481 | 1.86 (0.21) | 1.85 (0.21) | 0.742 |
| **Creatinine (μmol/L)** | 72.12 (14.09) | 72.12 (14.09) | 72.36 (14.19) | 0.836 | 72.11 (14.09) | 72.85 (14.73) | 0.461 |
| **Dead, n (%)** | 221 (1.2) | 221 (1.2) | 0 (0.0) | 0.322 | 221 (1.2) | 0 (0.0) | 0.199 |
| **Age at death, yrs** | 70.73 (6.71) | 70.73 (6.71) | NaN (NA) | NA | 70.73 (6.71) | NaN (NA) | NA |
| **Coronary artery disease, n (%)** | 846 (4.5) | 838 (4.5) | 8 (5.2) | 0.84 | 831 (4.5) | 15 (7.1) | 0.104 |
| **Heart failure, n (%)** | 240 (1.3) | 233 (1.3) | 7 (4.5) | 0.001 | 230 (1.2) | 10 (4.7) | <0.001 |
| **Early DCM features, n (%)** |  |  |  | 0.0019 |  |  | 0.0026 |
| **Arrhythmia and/or cardiac conduction disease** | 2729 (14.6) | 2710 (14.6) | 19 (12.3) |  | 2696 (14.6) | 33 (15.6) |  |
| **Isolated ventricular dilatation** | 522 (2.8) | 518 (2.8) | 4 (2.6) |  | 518 (2.8) | 4 (1.9) |  |
| **Hypokinetic non-dilated cardiomyopathy** | 645 (3.5) | 631 (3.4) | 14 (9.1) |  | 628 (3.4) | 17 (8.0) |  |
| **DCM overall, n (%)** | 205 (1.1) | 202 (1.1) | 3 (1.9) | 0.31 | 200 (1.1) | 5 (2.4) | 0.077 |
| **Subclinical DCM** | 189 (1.0) | 188 (1.0) | 1 (0.6) |  | 187 (1.0) | 2 (0.9) |  |
| **Clinical DCM** | 19 (0.1) | 17 (0.1) | 2 (1.3) |  | 16 (0.1) | 3 (1.4) |  |
| **LVESV (ml)** | 63.46 (63.75) | 63.42 (63.76) | 67.88 (63.52) | 0.387 | 63.42 (63.84) | 66.62 (56.02) | 0.467 |
| **LVEDV (ml)** | 140.72 (136.83) | 140.71 (137.24) | 142.75 (73.48) | 0.854 | 140.72 (137.44) | 141.23 (65.53) | 0.957 |
| **LVEF (%)** | 55.67 (6.62) | 55.68 (6.60) | 54.50 (8.43) | 0.028 | 55.68 (6.60) | 54.55 (8.05) | 0.013 |
| **Hypertension, n (%)** | 6028 (32.3) | 5981 (32.3) | 47 (30.5) | 0.699 | 5961 (32.3) | 67 (31.6) | 0.886 |
| **Stroke, n (%)** | 330 (1.8) | 329 (1.8) | 1 (0.6) | 0.453 | 328 (1.8) | 2 (0.9) | 0.513 |
| **Valvular heart disease, n (%)** | 527 (2.8) | 523 (2.8) | 4 (2.6) | 1 | 519 (2.8) | 8 (3.8) | 0.528 |
| **Atrial fibrillation/flutter, n (%)** | 832 (4.5) | 822 (4.4) | 10 (6.5) | 0.301 | 818 (4.4) | 14 (6.6) | 0.175 |
| **Bradyarrhythmia, n (%)** | 352 (1.9) | 350 (1.9) | 2 (1.3) | 0.81 | 349 (1.9) | 3 (1.4) | 0.8 |
| **Cardiac conduction defect, n (%)** | 2448 (13.1) | 2432 (13.1) | 16 (10.4) | 0.375 | 2423 (13.1) | 25 (11.8) | 0.637 |
| **Sick sinus syndrome, n (%)** | 19 (0.1) | 19 (0.1) | 0 (0.0) | 1 | 19 (0.1) | 0 (0.0) | 1 |
| **CIED, n (%)** | 148 (0.8) | 147 (0.8) | 1 (0.6) | 1 | 146 (0.8) | 2 (0.9) | 1 |
| **Pre-excitation syndrome, n (%)** | 35 (0.2) | 35 (0.2) | 0 (0.0) | 1 | 33 (0.2) | 2 (0.9) | 0.078 |
| **Ventricular arrhythmia, n (%)** | 69 (0.4) | 68 (0.4) | 1 (0.6) | 1 | 67 (0.4) | 2 (0.9) | 0.415 |
| **eGFR (ml/min/1.73m^2^)** | 87.32 (16.24) | 87.32 (16.26) | 86.98 (14.27) | 0.801 | 87.32 (16.26) | 87.23 (14.64) | 0.936 |
| **CKD ≥3, n (%)** | 396 (2.2) | 392 (2.2) | 4 (2.7) | 0.907 | 390 (2.2) | 6 (3.0) | 0.615 |

BP = blood pressure; DCM = dilated cardiomyopathy; CIED = cardiac implantable electronic device (including single- and dual-chamber permanent pacemaker, implantable cardioverter defibrillator, cardiac resynchronization therapy); CKD = chronic kidney disease; eGFR = estimated glomerular filtration rate; LVEDV = left ventricular end diastolic volume; LVESV = left ventricular end systolic volume; LVEF = left ventricular ejection fraction.

### Table S4. Clinical and subclinical penetrance of DCM and DCM-associated clinical features in participants with putative pathogenic variants in DCM genes.

|  | Filter | Overall | Phenotype negative | Phenotype positive | | | |
| --- | --- | --- | --- | --- | --- | --- | --- |
|  |  |  |  | Early DCM features | | | DCM |
|  |  |  |  | Arrhythmia and/or cardiac conduction disease | Isolated ventricular dilatation | Hypokinetic  non-dilated cardiomyopathy |  |
| Clinical | Total | 18665 | 17545 (94%) | 1101 (5.9%) | NA | NA | 19 (0.1%) |
|  | missense pLOF FAF | 1463 (7.8%) | 1374 (93.92%) | 84 (5.74%) | NA | NA | 5 (0.34%) |
|  | InterVar FAF | 154 (0.8%) | 142 (92.21%) | 10 (6.49%) | NA | NA | 2 (1.3%) |
|  | InterVar FAF ClinVar 2* | 212 (1.1%) | 193 (91.04%) | 16 (7.55%) | NA | NA | 3 (1.42%) |
| Subclinical phenotype (MRI + ECG) | Total | 18665 | 15056 (80.66%) | 2253 (12.07%) | 522 (2.8%) | 645 (3.46%) | 189 (1.01%) |
|  | missense pLOF FAF | 1463 (7.8%) | 1147 (78.4%) | 194 (13.26%) | 35 (2.39%) | 63 (4.31%) | 24 (1.64%) |
|  | InterVar FAF | 154 (0.8%) | 121 (78.57%) | 14 (9.09%) | 4 (2.6%) | 14 (9.09%) | 1 (0.65%) |
|  | InterVar FAF ClinVar 2* | 212 (1.1%) | 165 (77.83%) | 24 (11.32%) | 4 (1.89%) | 17 (8.02%) | 2 (0.94%) |
| Combined | Total | 18665 | 14578 (78.1%) | 2725 (14.6%) | 522 (2.8%) | 635 (3.4%) | 205 (1.1%) |
|  | missense pLOF FAF | 1463 (7.8%) | 1117 (76.35%) | 223 (15.24%) | 35 (2.39%) | 59 (4.03%) | 29 (1.98%) |
|  | InterVar FAF | 154 (0.8%) | 116 (75.32%) | 19 (12.34%) | 4 (2.6%) | 12 (7.79%) | 3 (1.95%) |
|  | InterVar FAF ClinVar 2* | 212 (1.1%) | 156 (73.58%) | 33 (15.57%) | 4 (1.89%) | 14 (6.61%) | 5 (2.36%) |

The number and proportion for each phenotype is shown for those with a putative pathogenic variant by each of the three filtering strategies and in the total study population for comparison.

**Table S5. Penetrance of putative pathogenic variants identified by missense pLOF FAF filtering strategy**

| Gene | Total | Phenotype negative | Arrhythmia and/or cardiac conduction disease | Isolated ventricular dilatation | Hypokinetic | DCM (CMR) |
| --- | --- | --- | --- | --- | --- | --- |
| *BAG3* | 5 (100%) | 5 (100%) | 0 (0%) | 0 (0%) | 0 (0%) | 0 (0%) |
| *DES* | 32 (100%) | 22 (68.75%) | 4 (12.5%) | 3 (9.38%) | 2 (6.25%) | 1 (3.12%) |
| *FLNC* | 121 (100%) | 93 (76.86%) | 21 (17.36%) | 4 (3.31%) | 1 (0.83%) | 2 (1.65%) |
| *LMNA* | 30 (100%) | 21 (70%) | 7 (23.33%) | 0 (0%) | 1 (3.33%) | 1 (3.33%) |
| *MYH7* | 122 (100%) | 100 (81.97%) | 16 (13.11%) | 0 (0%) | 3 (2.46%) | 3 (2.46%) |
| *PLN* | 5 (100%) | 5 (100%) | 0 (0%) | 0 (0%) | 0 (0%) | 0 (0%) |
| *RBM20* | 8 (100%) | 7 (87.5%) | 1 (12.5%) | 0 (0%) | 0 (0%) | 0 (0%) |
| *SCN5A* | 140 (100%) | 115 (82.14%) | 16 (11.43%) | 2 (1.43%) | 4 (2.86%) | 3 (2.14%) |
| *TNNC1* | 4 (100%) | 3 (75%) | 1 (25%) | 0 (0%) | 0 (0%) | 0 (0%) |
| *TNNT2* | 19 (100%) | 17 (89.47%) | 2 (10.53%) | 0 (0%) | 0 (0%) | 0 (0%) |
| *TTN* | 44 (100%) | 22 (50%) | 5 (11.36%) | 2 (4.55%) | 15 (34.09%) | 0 (0%) |
| *DSP* | 43 (100%) | 33 (76.74%) | 6 (13.95%) | 0 (0%) | 3 (6.98%) | 1 (2.33%) |
| *ACTC1* | 6 (100%) | 5 (83.33%) | 0 (0%) | 1 (16.67%) | 0 (0%) | 0 (0%) |
| *ACTN2* | 26 (100%) | 16 (61.54%) | 7 (26.92%) | 0 (0%) | 0 (0%) | 3 (11.54%) |
| *JPH2* | 5 (100%) | 4 (80%) | 1 (20%) | 0 (0%) | 0 (0%) | 0 (0%) |
| *NEXN* | 17 (100%) | 14 (82.35%) | 0 (0%) | 0 (0%) | 3 (17.65%) | 0 (0%) |
| *TNNI3* | 4 (100%) | 3 (75%) | 1 (25%) | 0 (0%) | 0 (0%) | 0 (0%) |
| *TPM1* | 9 (100%) | 8 (88.89%) | 1 (11.11%) | 0 (0%) | 0 (0%) | 0 (0%) |
| *VCL* | 11 (100%) | 10 (90.91%) | 1 (9.09%) | 0 (0%) | 0 (0%) | 0 (0%) |
| *ABCC9* | 44 (100%) | 31 (70.45%) | 11 (25%) | 1 (2.27%) | 0 (0%) | 1 (2.27%) |
| *ANKRD1* | 3 (100%) | 1 (33.33%) | 1 (33.33%) | 0 (0%) | 1 (33.33%) | 0 (0%) |
| *CSRP3* | 21 (100%) | 16 (76.19%) | 2 (9.52%) | 1 (4.76%) | 2 (9.52%) | 0 (0%) |
| *CTF1* | 3 (100%) | 3 (100%) | 0 (0%) | 0 (0%) | 0 (0%) | 0 (0%) |
| *DSG2* | 25 (100%) | 21 (84%) | 2 (8%) | 1 (4%) | 0 (0%) | 1 (4%) |
| *DTNA* | 9 (100%) | 8 (88.89%) | 1 (11.11%) | 0 (0%) | 0 (0%) | 0 (0%) |
| *EYA4* | 27 (100%) | 19 (70.37%) | 7 (25.93%) | 1 (3.7%) | 0 (0%) | 0 (0%) |
| *GATAD1* | 6 (100%) | 5 (83.33%) | 0 (0%) | 1 (16.67%) | 0 (0%) | 0 (0%) |
| *ILK* | 28 (100%) | 23 (82.14%) | 3 (10.71%) | 1 (3.57%) | 1 (3.57%) | 0 (0%) |
| *LAMA4* | 27 (100%) | 14 (51.85%) | 12 (44.44%) | 0 (0%) | 0 (0%) | 1 (3.7%) |
| *LDB3* | 27 (100%) | 16 (59.26%) | 9 (33.33%) | 1 (3.7%) | 1 (3.7%) | 0 (0%) |
| *MYBPC3* | 46 (100%) | 31 (67.39%) | 10 (21.74%) | 3 (6.52%) | 2 (4.35%) | 0 (0%) |
| *MYH6* | 149 (100%) | 114 (76.51%) | 24 (16.11%) | 4 (2.68%) | 4 (2.68%) | 3 (2.01%) |
| *MYL2* | 14 (100%) | 10 (71.43%) | 3 (21.43%) | 0 (0%) | 1 (7.14%) | 0 (0%) |
| *MYPN* | 15 (100%) | 11 (73.33%) | 3 (20%) | 0 (0%) | 1 (6.67%) | 0 (0%) |
| *NEBL* | 26 (100%) | 21 (80.77%) | 4 (15.38%) | 1 (3.85%) | 0 (0%) | 0 (0%) |
| *NKX2-5* | 3 (100%) | 3 (100%) | 0 (0%) | 0 (0%) | 0 (0%) | 0 (0%) |
| *OBSCN* | 153 (100%) | 122 (79.74%) | 16 (10.46%) | 4 (2.61%) | 10 (6.54%) | 1 (0.65%) |
| *PLEKHM2* | 8 (100%) | 5 (62.5%) | 3 (37.5%) | 0 (0%) | 0 (0%) | 0 (0%) |
| *PRDM16* | 2 (100%) | 2 (100%) | 0 (0%) | 0 (0%) | 0 (0%) | 0 (0%) |
| *PSEN2* | 36 (100%) | 31 (86.11%) | 2 (5.56%) | 0 (0%) | 1 (2.78%) | 2 (5.56%) |
| *SGCD* | 16 (100%) | 12 (75%) | 2 (12.5%) | 1 (6.25%) | 1 (6.25%) | 0 (0%) |
| *TBX20* | 7 (100%) | 6 (85.71%) | 1 (14.29%) | 0 (0%) | 0 (0%) | 0 (0%) |
| *TCAP* | 8 (100%) | 6 (75%) | 1 (12.5%) | 0 (0%) | 1 (12.5%) | 0 (0%) |
| *TNNI3K* | 41 (100%) | 34 (82.93%) | 5 (12.2%) | 1 (2.44%) | 1 (2.44%) | 0 (0%) |
| *ABCC9, FLNC* | 1 (100%) | 1 (100%) | 0 (0%) | 0 (0%) | 0 (0%) | 0 (0%) |
| *ABCC9, MYH6* | 1 (100%) | 1 (100%) | 0 (0%) | 0 (0%) | 0 (0%) | 0 (0%) |
| *ACTN2, MYH6* | 1 (100%) | 1 (100%) | 0 (0%) | 0 (0%) | 0 (0%) | 0 (0%) |
| *ACTN2, TTN* | 1 (100%) | 1 (100%) | 0 (0%) | 0 (0%) | 0 (0%) | 0 (0%) |
| *CSRP3, SCN5A* | 1 (100%) | 1 (100%) | 0 (0%) | 0 (0%) | 0 (0%) | 0 (0%) |
| *CTF1, MYH6* | 1 (100%) | 1 (100%) | 0 (0%) | 0 (0%) | 0 (0%) | 0 (0%) |
| *DES, LAMA4* | 1 (100%) | 1 (100%) | 0 (0%) | 0 (0%) | 0 (0%) | 0 (0%) |
| *DES, MYBPC3* | 1 (100%) | 1 (100%) | 0 (0%) | 0 (0%) | 0 (0%) | 0 (0%) |
| *DES, MYH6* | 1 (100%) | 0 (0%) | 1 (100%) | 0 (0%) | 0 (0%) | 0 (0%) |
| *DES, SCN5A* | 1 (100%) | 1 (100%) | 0 (0%) | 0 (0%) | 0 (0%) | 0 (0%) |
| *DES, TNNI3* | 1 (100%) | 0 (0%) | 0 (0%) | 0 (0%) | 1 (100%) | 0 (0%) |
| *DSG2, OBSCN* | 1 (100%) | 0 (0%) | 0 (0%) | 1 (100%) | 0 (0%) | 0 (0%) |
| *DSP, EYA4* | 1 (100%) | 1 (100%) | 0 (0%) | 0 (0%) | 0 (0%) | 0 (0%) |
| *DSP, MYH6* | 1 (100%) | 1 (100%) | 0 (0%) | 0 (0%) | 0 (0%) | 0 (0%) |
| *DSP, MYH7, TNNI3K* | 1 (100%) | 1 (100%) | 0 (0%) | 0 (0%) | 0 (0%) | 0 (0%) |
| *DSP, SCN5A* | 1 (100%) | 1 (100%) | 0 (0%) | 0 (0%) | 0 (0%) | 0 (0%) |
| *DTNA, NEXN* | 1 (100%) | 0 (0%) | 1 (100%) | 0 (0%) | 0 (0%) | 0 (0%) |
| *EYA4, EYA4* | 1 (100%) | 1 (100%) | 0 (0%) | 0 (0%) | 0 (0%) | 0 (0%) |
| *EYA4, LAMA4* | 1 (100%) | 1 (100%) | 0 (0%) | 0 (0%) | 0 (0%) | 0 (0%) |
| *EYA4, MYH6* | 2 (100%) | 0 (0%) | 2 (100%) | 0 (0%) | 0 (0%) | 0 (0%) |
| *EYA4, MYH7* | 1 (100%) | 1 (100%) | 0 (0%) | 0 (0%) | 0 (0%) | 0 (0%) |
| *FLNC, FLNC* | 1 (100%) | 1 (100%) | 0 (0%) | 0 (0%) | 0 (0%) | 0 (0%) |
| *FLNC, MYBPC3* | 2 (100%) | 2 (100%) | 0 (0%) | 0 (0%) | 0 (0%) | 0 (0%) |
| *FLNC, MYH7* | 2 (100%) | 1 (50%) | 0 (0%) | 0 (0%) | 1 (50%) | 0 (0%) |
| *FLNC, MYL2* | 1 (100%) | 0 (0%) | 1 (100%) | 0 (0%) | 0 (0%) | 0 (0%) |
| *FLNC, OBSCN* | 1 (100%) | 1 (100%) | 0 (0%) | 0 (0%) | 0 (0%) | 0 (0%) |
| *FLNC, PSEN2* | 1 (100%) | 1 (100%) | 0 (0%) | 0 (0%) | 0 (0%) | 0 (0%) |
| *FLNC, TNNT2* | 1 (100%) | 0 (0%) | 0 (0%) | 1 (100%) | 0 (0%) | 0 (0%) |
| *LAMA4, LAMA4* | 1 (100%) | 1 (100%) | 0 (0%) | 0 (0%) | 0 (0%) | 0 (0%) |
| *LAMA4, LDB3* | 1 (100%) | 1 (100%) | 0 (0%) | 0 (0%) | 0 (0%) | 0 (0%) |
| *LAMA4, SCN5A* | 1 (100%) | 0 (0%) | 1 (100%) | 0 (0%) | 0 (0%) | 0 (0%) |
| *LMNA, MYBPC3, MYH7* | 1 (100%) | 0 (0%) | 0 (0%) | 0 (0%) | 1 (100%) | 0 (0%) |
| *MYH6, MYH6* | 2 (100%) | 1 (50%) | 0 (0%) | 0 (0%) | 1 (50%) | 0 (0%) |
| *MYH6, MYH7* | 1 (100%) | 1 (100%) | 0 (0%) | 0 (0%) | 0 (0%) | 0 (0%) |
| *MYH6, MYL2* | 1 (100%) | 1 (100%) | 0 (0%) | 0 (0%) | 0 (0%) | 0 (0%) |
| *MYH6, OBSCN* | 4 (100%) | 4 (100%) | 0 (0%) | 0 (0%) | 0 (0%) | 0 (0%) |
| *MYH6, RBM20* | 1 (100%) | 1 (100%) | 0 (0%) | 0 (0%) | 0 (0%) | 0 (0%) |
| *MYH6, SCN5A* | 3 (100%) | 2 (66.67%) | 1 (33.33%) | 0 (0%) | 0 (0%) | 0 (0%) |
| *MYH7, MYH7, MYH7* | 1 (100%) | 1 (100%) | 0 (0%) | 0 (0%) | 0 (0%) | 0 (0%) |
| *MYH7, RBM20* | 1 (100%) | 1 (100%) | 0 (0%) | 0 (0%) | 0 (0%) | 0 (0%) |
| *MYH7, SCN5A* | 1 (100%) | 0 (0%) | 1 (100%) | 0 (0%) | 0 (0%) | 0 (0%) |
| *MYL2, SCN5A* | 1 (100%) | 0 (0%) | 1 (100%) | 0 (0%) | 0 (0%) | 0 (0%) |
| *NEXN, NEXN* | 1 (100%) | 1 (100%) | 0 (0%) | 0 (0%) | 0 (0%) | 0 (0%) |
| *NEXN, TTN* | 1 (100%) | 1 (100%) | 0 (0%) | 0 (0%) | 0 (0%) | 0 (0%) |
| *OBSCN, OBSCN* | 5 (100%) | 5 (100%) | 0 (0%) | 0 (0%) | 0 (0%) | 0 (0%) |
| *OBSCN, PSEN2* | 1 (100%) | 0 (0%) | 0 (0%) | 0 (0%) | 0 (0%) | 1 (100%) |
| *OBSCN, SCN5A* | 1 (100%) | 1 (100%) | 0 (0%) | 0 (0%) | 0 (0%) | 0 (0%) |
| *OBSCN, TCAP* | 1 (100%) | 1 (100%) | 0 (0%) | 0 (0%) | 0 (0%) | 0 (0%) |
| *PSEN2, TNNI3K* | 1 (100%) | 0 (0%) | 1 (100%) | 0 (0%) | 0 (0%) | 0 (0%) |
| *SCN5A, SCN5A* | 1 (100%) | 0 (0%) | 1 (100%) | 0 (0%) | 0 (0%) | 0 (0%) |
| *TBX20, TNNI3K* | 1 (100%) | 0 (0%) | 1 (100%) | 0 (0%) | 0 (0%) | 0 (0%) |
| *TNNI3K, TNNI3K* | 1 (100%) | 1 (100%) | 0 (0%) | 0 (0%) | 0 (0%) | 0 (0%) |
| *TPM1, TPM1* | 1 (100%) | 1 (100%) | 0 (0%) | 0 (0%) | 0 (0%) | 0 (0%) |
| *TTN, TTN* | 2 (100%) | 2 (100%) | 0 (0%) | 0 (0%) | 0 (0%) | 0 (0%) |

**Table S6. Penetrance of putative pathogenic variants identified by InterVar FAF filtering strategy.**

| Gene | Total | Phenotype negative | Arrhythmia and/or cardiac conduction disease | Isolated ventricular dilatation | Hypokinetic | DCM |
| --- | --- | --- | --- | --- | --- | --- |
| *DES* | 4 (100%) | 3 (75%) | 0 (0%) | 0 (0%) | 1 (25%) | 0 (0%) |
| *FLNC* | 14 (100%) | 10 (71.43%) | 3 (21.43%) | 1 (7.14%) | 0 (0%) | 0 (0%) |
| *LMNA* | 1 (100%) | 1 (100%) | 0 (0%) | 0 (0%) | 0 (0%) | 0 (0%) |
| *PLN* | 1 (100%) | 1 (100%) | 0 (0%) | 0 (0%) | 0 (0%) | 0 (0%) |
| *SCN5A* | 13 (100%) | 11 (84.62%) | 0 (0%) | 0 (0%) | 1 (7.69%) | 1 (7.69%) |
| *TNNC1* | 6 (100%) | 6 (100%) | 0 (0%) | 0 (0%) | 0 (0%) | 0 (0%) |
| *TTN* | 25 (100%) | 16 (64%) | 2 (8%) | 1 (4%) | 6 (24%) | 0 (0%) |
| *DSP* | 7 (100%) | 6 (85.71%) | 0 (0%) | 0 (0%) | 1 (14.29%) | 0 (0%) |
| *ACTN2* | 1 (100%) | 0 (0%) | 1 (100%) | 0 (0%) | 0 (0%) | 0 (0%) |
| *JPH2* | 1 (100%) | 1 (100%) | 0 (0%) | 0 (0%) | 0 (0%) | 0 (0%) |
| *NEXN* | 4 (100%) | 4 (100%) | 0 (0%) | 0 (0%) | 0 (0%) | 0 (0%) |
| *TPM1* | 1 (100%) | 1 (100%) | 0 (0%) | 0 (0%) | 0 (0%) | 0 (0%) |
| *ABCC9* | 6 (100%) | 4 (66.67%) | 1 (16.67%) | 1 (16.67%) | 0 (0%) | 0 (0%) |
| *CSRP3* | 2 (100%) | 2 (100%) | 0 (0%) | 0 (0%) | 0 (0%) | 0 (0%) |
| *DSG2* | 2 (100%) | 1 (50%) | 0 (0%) | 1 (50%) | 0 (0%) | 0 (0%) |
| *EYA4* | 1 (100%) | 1 (100%) | 0 (0%) | 0 (0%) | 0 (0%) | 0 (0%) |
| *LAMA4* | 5 (100%) | 3 (60%) | 2 (40%) | 0 (0%) | 0 (0%) | 0 (0%) |
| *LDB3* | 4 (100%) | 3 (75%) | 1 (25%) | 0 (0%) | 0 (0%) | 0 (0%) |
| *MYBPC3* | 1 (100%) | 1 (100%) | 0 (0%) | 0 (0%) | 0 (0%) | 0 (0%) |
| *MYH6* | 2 (100%) | 2 (100%) | 0 (0%) | 0 (0%) | 0 (0%) | 0 (0%) |
| *MYL2* | 3 (100%) | 2 (66.67%) | 1 (33.33%) | 0 (0%) | 0 (0%) | 0 (0%) |
| *MYPN* | 3 (100%) | 2 (66.67%) | 1 (33.33%) | 0 (0%) | 0 (0%) | 0 (0%) |
| *NEBL* | 7 (100%) | 6 (85.71%) | 1 (14.29%) | 0 (0%) | 0 (0%) | 0 (0%) |
| *OBSCN* | 31 (100%) | 22 (70.97%) | 4 (12.9%) | 0 (0%) | 5 (16.13%) | 0 (0%) |
| *SGCD* | 1 (100%) | 1 (100%) | 0 (0%) | 0 (0%) | 0 (0%) | 0 (0%) |
| *TCAP* | 2 (100%) | 1 (50%) | 1 (50%) | 0 (0%) | 0 (0%) | 0 (0%) |
| *TNNI3K* | 6 (100%) | 5 (83.33%) | 1 (16.67%) | 0 (0%) | 0 (0%) | 0 (0%) |

**Table S7. Penetrance of putative pathogenic variants identified by InterVar FAF ClinVar filtering strategy**

|  | Total | Phenotype negative | Arrhythmia and/or cardiac conduction disease | Isolated ventricular dilatation | Hypokinetic non-dilated cardiomyopathy | DCM |
| --- | --- | --- | --- | --- | --- | --- |
| *DES* | 4 (100%) | 3 (75%) | 0 (0%) | 0 (0%) | 1 (25%) | 0 (0%) |
| *FLNC* | 16 (100%) | 11 (68.75%) | 4 (25%) | 1 (6.25%) | 0 (0%) | 0 (0%) |
| *LMNA* | 1 (100%) | 1 (100%) | 0 (0%) | 0 (0%) | 0 (0%) | 0 (0%) |
| *MYH7* | 9 (100%) | 8 (88.89%) | 0 (0%) | 0 (0%) | 0 (0%) | 1 (11.11%) |
| *PLN* | 1 (100%) | 1 (100%) | 0 (0%) | 0 (0%) | 0 (0%) | 0 (0%) |
| *SCN5A* | 18 (100%) | 12 (66.67%) | 4 (22.22%) | 0 (0%) | 1 (5.56%) | 1 (5.56%) |
| *TNNC1* | 6 (100%) | 6 (100%) | 0 (0%) | 0 (0%) | 0 (0%) | 0 (0%) |
| *TTN* | 35 (100%) | 20 (57.14%) | 6 (17.14%) | 1 (2.86%) | 8 (22.86%) | 0 (0%) |
| *DSP* | 8 (100%) | 6 (75%) | 0 (0%) | 0 (0%) | 2 (25%) | 0 (0%) |
| *ACTN2* | 1 (100%) | 0 (0%) | 1 (100%) | 0 (0%) | 0 (0%) | 0 (0%) |
| *JPH2* | 1 (100%) | 1 (100%) | 0 (0%) | 0 (0%) | 0 (0%) | 0 (0%) |
| *NEXN* | 4 (100%) | 4 (100%) | 0 (0%) | 0 (0%) | 0 (0%) | 0 (0%) |
| *TNNI3* | 2 (100%) | 2 (100%) | 0 (0%) | 0 (0%) | 0 (0%) | 0 (0%) |
| *TPM1* | 1 (100%) | 1 (100%) | 0 (0%) | 0 (0%) | 0 (0%) | 0 (0%) |
| *ABCC9* | 6 (100%) | 4 (66.67%) | 1 (16.67%) | 1 (16.67%) | 0 (0%) | 0 (0%) |
| *CSRP3* | 2 (100%) | 2 (100%) | 0 (0%) | 0 (0%) | 0 (0%) | 0 (0%) |
| *DSG2* | 9 (100%) | 7 (77.78%) | 1 (11.11%) | 1 (11.11%) | 0 (0%) | 0 (0%) |
| *EYA4* | 1 (100%) | 1 (100%) | 0 (0%) | 0 (0%) | 0 (0%) | 0 (0%) |
| *LAMA4* | 5 (100%) | 3 (60%) | 2 (40%) | 0 (0%) | 0 (0%) | 0 (0%) |
| *LDB3* | 4 (100%) | 3 (75%) | 1 (25%) | 0 (0%) | 0 (0%) | 0 (0%) |
| *MYBPC3* | 22 (100%) | 18 (81.82%) | 4 (18.18%) | 0 (0%) | 0 (0%) | 0 (0%) |
| *MYH6* | 2 (100%) | 2 (100%) | 0 (0%) | 0 (0%) | 0 (0%) | 0 (0%) |
| *MYL2* | 4 (100%) | 3 (75%) | 1 (25%) | 0 (0%) | 0 (0%) | 0 (0%) |
| *MYPN* | 3 (100%) | 2 (66.67%) | 1 (33.33%) | 0 (0%) | 0 (0%) | 0 (0%) |
| *NEBL* | 7 (100%) | 6 (85.71%) | 1 (14.29%) | 0 (0%) | 0 (0%) | 0 (0%) |
| *OBSCN* | 31 (100%) | 22 (70.97%) | 4 (12.9%) | 0 (0%) | 5 (16.13%) | 0 (0%) |
| *SGCD* | 1 (100%) | 1 (100%) | 0 (0%) | 0 (0%) | 0 (0%) | 0 (0%) |
| *TCAP* | 2 (100%) | 1 (50%) | 1 (50%) | 0 (0%) | 0 (0%) | 0 (0%) |
| *TNNI3K* | 6 (100%) | 5 (83.33%) | 1 (16.67%) | 0 (0%) | 0 (0%) | 0 (0%) |

**Table S8. Incidence and prevalence of DCM and DCM-associated features in overall UK Biobank population.**

|  |  |  | Prevalence | | Incidence | |
| --- | --- | --- | --- | --- | --- | --- |
|  | Filter |  | N | Prevalence (CI) [%] | Events/Person-Time | Incidence per 1000 person-years (CI) |
| AF | missense pLOF FAF | G- | 2526 | 1.37 (1.32 - 1.43 ) | 9592/2169629 | 4.42 (4.33 - 4.51 ) |
|  |  | G+ | 281 | 1.69 (1.5 - 1.89 ) | 970/194817 | 4.98 (4.67 - 5.3 ) |
|  |  | Unknown | 4515 | 1.5 (1.45 - 1.54 ) | 17204/3605810 | 4.77 (4.7 - 4.84 ) |
|  | InterVar FAF | G- | 2767 | 1.39 (1.34 - 1.44 ) | 10436/2344765 | 4.45 (4.37 - 4.54 ) |
|  |  | G+ | 40 | 2.36 (1.69 - 3.19 ) | 126/19681 | 6.4 (5.33 - 7.62 ) |
|  |  | Unknown | 4515 | 1.5 (1.45 - 1.54 ) | 17204/3605810 | 4.77 (4.7 - 4.84 ) |
|  | InterVar FAF ClinVar 2* | G- | 2751 | 1.39 (1.34 - 1.44 ) | 10377/2337910 | 4.44 (4.35 - 4.52 ) |
|  |  | G+ | 56 | 2.44 (1.85 - 3.15 ) | 185/26536 | 6.97 (6 - 8.05 ) |
|  |  | Unknown | 4515 | 1.5 (1.45 - 1.54 ) | 17204/3605810 | 4.77 (4.7 - 4.84 ) |
|  | Total | NA | 7322 | 1.46 (1.42 - 1.49 ) | 27766/5970255 | 4.65 (4.6 - 4.71 ) |
| Bradyarrhythmia | missense pLOF FAF | G- | 2352 | 1.28 (1.23 - 1.33 ) | 3641/2198600 | 1.66 (1.6 - 1.71 ) |
|  |  | G+ | 244 | 1.46 (1.29 - 1.66 ) | 358/197951 | 1.81 (1.63 - 2.01 ) |
|  |  | Unknown | 4399 | 1.46 (1.41 - 1.5 ) | 6720/3654672 | 1.84 (1.8 - 1.88 ) |
|  | InterVar FAF | G- | 2571 | 1.29 (1.24 - 1.34 ) | 3962/2376344 | 1.67 (1.62 - 1.72 ) |
|  |  | G+ | 25 | 1.47 (0.955 - 2.17 ) | 37/20207 | 1.83 (1.29 - 2.52 ) |
|  |  | Unknown | 4399 | 1.46 (1.41 - 1.5 ) | 6720/3654672 | 1.84 (1.8 - 1.88 ) |
|  | InterVar FAF ClinVar 2* | G- | 2554 | 1.29 (1.24 - 1.34 ) | 3938/2369370 | 1.66 (1.61 - 1.71 ) |
|  |  | G+ | 42 | 1.83 (1.32 - 2.46 ) | 61/27181 | 2.24 (1.72 - 2.88 ) |
|  |  | Unknown | 4399 | 1.46 (1.41 - 1.5 ) | 6720/3654672 | 1.84 (1.8 - 1.88 ) |
|  | Total | NA | 6995 | 1.39 (1.36 - 1.42 ) | 10719/6051223 | 1.77 (1.74 - 1.81 ) |
| Conduction Defect | missense pLOF FAF | G- | 2708 | 1.47 (1.42 - 1.53 ) | 1723/2202347 | 0.782 (0.746 - 0.82 ) |
|  |  | G+ | 270 | 1.62 (1.43 - 1.82 ) | 167/198560 | 0.841 (0.718 - 0.979 ) |
|  |  | Unknown | 5144 | 1.7 (1.66 - 1.75 ) | 3147/3661399 | 0.86 (0.83 - 0.89 ) |
|  | InterVar FAF | G- | 2946 | 1.48 (1.43 - 1.54 ) | 1871/2380662 | 0.786 (0.751 - 0.822 ) |
|  |  | G+ | 32 | 1.88 (1.29 - 2.65 ) | 19/20246 | 0.938 (0.565 - 1.47 ) |
|  |  | Unknown | 5144 | 1.7 (1.66 - 1.75 ) | 3147/3661399 | 0.86 (0.83 - 0.89 ) |
|  | InterVar FAF ClinVar 2* | G- | 2926 | 1.48 (1.42 - 1.53 ) | 1866/2373648 | 0.786 (0.751 - 0.823 ) |
|  |  | G+ | 52 | 2.26 (1.69 - 2.96 ) | 24/27260 | 0.88 (0.564 - 1.31 ) |
|  |  | Unknown | 5144 | 1.7 (1.66 - 1.75 ) | 3147/3661399 | 0.86 (0.83 - 0.89 ) |
|  | Total | NA | 8122 | 1.62 (1.58 - 1.65 ) | 5037/6062306 | 0.831 (0.808 - 0.854 ) |
| Sick sinus syndrome | missense pLOF FAF | G- | 414 | 0.225 (0.204 - 0.248 ) | 2/2235953 | 0.000894 (0.000108 - 0.00323 ) |
|  |  | G+ | 46 | 0.276 (0.202 - 0.368 ) | 0/201789 | 0 (0 - 0.0183 ) |
|  |  | Unknown | 761 | 0.252 (0.235 - 0.271 ) | 2/3725456 | 0.000537 (6.5e-05 - 0.00194 ) |
|  | InterVar FAF | G- | 455 | 0.229 (0.208 - 0.251 ) | 2/2417136 | 0.000827 (1e-04 - 0.00299 ) |
|  |  | G+ | 5 | 0.294 (0.0957 - 0.686 ) | 0/20606 | 0 (0 - 0.179 ) |
|  |  | Unknown | 761 | 0.252 (0.235 - 0.271 ) | 2/3725456 | 0.000537 (6.5e-05 - 0.00194 ) |
|  | InterVar FAF ClinVar 2* | G- | 453 | 0.228 (0.208 - 0.25 ) | 2/2409907 | 0.00083 (0.000101 - 0.003 ) |
|  |  | G+ | 7 | 0.304 (0.123 - 0.626 ) | 0/27834 | 0 (0 - 0.133 ) |
|  |  | Unknown | 761 | 0.252 (0.235 - 0.271 ) | 2/3725456 | 0.000537 (6.5e-05 - 0.00194 ) |
|  | Total | NA | 1221 | 0.243 (0.23 - 0.257 ) | 4/6163198 | 0.000649 (0.000177 - 0.00166 ) |
| Pre-excitation syndrome | missense pLOF FAF | G- | 89 | 0.0484 (0.0389 - 0.0595 ) | 102/2239231 | 0.0456 (0.0371 - 0.0553 ) |
|  |  | G+ | 8 | 0.048 (0.0207 - 0.0945 ) | 9/202200 | 0.0445 (0.0204 - 0.0845 ) |
|  |  | Unknown | 130 | 0.0431 (0.036 - 0.0511 ) | 194/3731990 | 0.052 (0.0449 - 0.0598 ) |
|  | InterVar FAF | G- | 96 | 0.0483 (0.0391 - 0.0589 ) | 111/2420777 | 0.0459 (0.0377 - 0.0552 ) |
|  |  | G+ | 1 | 0.0589 (0.00149 - 0.328 ) | 0/20654 | 0 (0 - 0.179 ) |
|  |  | Unknown | 130 | 0.0431 (0.036 - 0.0511 ) | 194/3731990 | 0.052 (0.0449 - 0.0598 ) |
|  | InterVar FAF ClinVar 2* | G- | 95 | 0.0479 (0.0388 - 0.0586 ) | 110/2413536 | 0.0456 (0.0375 - 0.0549 ) |
|  |  | G+ | 2 | 0.087 (0.0105 - 0.314 ) | 1/27895 | 0.0358 (0.000908 - 0.2 ) |
|  |  | Unknown | 130 | 0.0431 (0.036 - 0.0511 ) | 194/3731990 | 0.052 (0.0449 - 0.0598 ) |
|  | Total | NA | 227 | 0.0452 (0.0395 - 0.0515 ) | 305/6173421 | 0.0494 (0.044 - 0.0553 ) |
| Ventricular arrhythmia | missense pLOF FAF | G- | 262 | 0.142 (0.126 - 0.161 ) | 1362/2233982 | 0.61 (0.578 - 0.643 ) |
|  |  | G+ | 28 | 0.168 (0.112 - 0.243 ) | 135/201627 | 0.67 (0.561 - 0.792 ) |
|  |  | Unknown | 623 | 0.206 (0.191 - 0.223 ) | 2680/3719562 | 0.721 (0.693 - 0.748 ) |
|  | InterVar FAF | G- | 284 | 0.143 (0.127 - 0.16 ) | 1474/2415072 | 0.61 (0.58 - 0.642 ) |
|  |  | G+ | 6 | 0.353 (0.13 - 0.768 ) | 23/20537 | 1.12 (0.71 - 1.68 ) |
|  |  | Unknown | 623 | 0.206 (0.191 - 0.223 ) | 2680/3719562 | 0.721 (0.693 - 0.748 ) |
|  | InterVar FAF ClinVar 2* | G- | 281 | 0.142 (0.126 - 0.159 ) | 1463/2407911 | 0.608 (0.577 - 0.64 ) |
|  |  | G+ | 9 | 0.391 (0.179 - 0.742 ) | 34/27698 | 1.23 (0.85 - 1.72 ) |
|  |  | Unknown | 623 | 0.206 (0.191 - 0.223 ) | 2680/3719562 | 0.721 (0.693 - 0.748 ) |
|  | Total | NA | 913 | 0.182 (0.17 - 0.194 ) | 4177/6155171 | 0.679 (0.658 - 0.7 ) |
| Heart failure | missense pLOF FAF | G- | 821 | 0.446 (0.416 - 0.478 ) | 4419/2216998 | 1.99 (1.93 - 2.05 ) |
|  |  | G+ | 114 | 0.684 (0.564 - 0.821 ) | 475/199410 | 2.38 (2.17 - 2.61 ) |
|  |  | Unknown | 1851 | 0.613 (0.586 - 0.642 ) | 9013/3682685 | 2.45 (2.4 - 2.5 ) |
|  | InterVar FAF | G- | 914 | 0.459 (0.43 - 0.49 ) | 4835/2396202 | 2.02 (1.96 - 2.08 ) |
|  |  | G+ | 21 | 1.24 (0.767 - 1.88 ) | 59/20206 | 2.92 (2.22 - 3.77 ) |
|  |  | Unknown | 1851 | 0.613 (0.586 - 0.642 ) | 9013/3682685 | 2.45 (2.4 - 2.5 ) |
|  | InterVar FAF ClinVar 2* | G- | 907 | 0.457 (0.428 - 0.488 ) | 4809/2389107 | 2.01 (1.96 - 2.07 ) |
|  |  | G+ | 28 | 1.22 (0.811 - 1.76 ) | 85/27300 | 3.11 (2.49 - 3.85 ) |
|  |  | Unknown | 1851 | 0.613 (0.586 - 0.642 ) | 9013/3682685 | 2.45 (2.4 - 2.5 ) |
|  | Total | NA | 2786 | 0.554 (0.534 - 0.575 ) | 13907/6099092 | 2.28 (2.24 - 2.32 ) |
| Stroke | missense pLOF FAF | G- | 2300 | 1.25 (1.2 - 1.3 ) | 2773/2202251 | 1.26 (1.21 - 1.31 ) |
|  |  | G+ | 218 | 1.31 (1.14 - 1.49 ) | 244/198846 | 1.23 (1.08 - 1.39 ) |
|  |  | Unknown | 4302 | 1.43 (1.38 - 1.47 ) | 4940/3663451 | 1.35 (1.31 - 1.39 ) |
|  | InterVar FAF | G- | 2493 | 1.25 (1.2 - 1.3 ) | 2995/2380803 | 1.26 (1.21 - 1.3 ) |
|  |  | G+ | 25 | 1.47 (0.955 - 2.17 ) | 22/20293 | 1.08 (0.679 - 1.64 ) |
|  |  | Unknown | 4302 | 1.43 (1.38 - 1.47 ) | 4940/3663451 | 1.35 (1.31 - 1.39 ) |
|  | InterVar FAF ClinVar 2* | G- | 2486 | 1.25 (1.21 - 1.3 ) | 2980/2373675 | 1.26 (1.21 - 1.3 ) |
|  |  | G+ | 32 | 1.39 (0.954 - 1.96 ) | 37/27421 | 1.35 (0.95 - 1.86 ) |
|  |  | Unknown | 4302 | 1.43 (1.38 - 1.47 ) | 4940/3663451 | 1.35 (1.31 - 1.39 ) |
|  | Total | NA | 6820 | 1.36 (1.33 - 1.39 ) | 7957/6064548 | 1.31 (1.28 - 1.34 ) |
| CIED | missense pLOF FAF | G- | 506 | 0.275 (0.252 - 0.3 ) | 2950/2222107 | 1.33 (1.28 - 1.38 ) |
|  |  | G+ | 61 | 0.366 (0.28 - 0.47 ) | 307/200190 | 1.53 (1.37 - 1.72 ) |
|  |  | Unknown | 1168 | 0.387 (0.365 - 0.41 ) | 5345/3697926 | 1.45 (1.41 - 1.48 ) |
|  | InterVar FAF | G- | 562 | 0.283 (0.26 - 0.307 ) | 3227/2401797 | 1.34 (1.3 - 1.39 ) |
|  |  | G+ | 5 | 0.294 (0.0957 - 0.686 ) | 30/20500 | 1.46 (0.987 - 2.09 ) |
|  |  | Unknown | 1168 | 0.387 (0.365 - 0.41 ) | 5345/3697926 | 1.45 (1.41 - 1.48 ) |
|  | InterVar FAF ClinVar 2* | G- | 552 | 0.278 (0.256 - 0.303 ) | 3212/2394739 | 1.34 (1.3 - 1.39 ) |
|  |  | G+ | 15 | 0.652 (0.366 - 1.07 ) | 45/27558 | 1.63 (1.19 - 2.19 ) |
|  |  | Unknown | 1168 | 0.387 (0.365 - 0.41 ) | 5345/3697926 | 1.45 (1.41 - 1.48 ) |
|  | Total | NA | 1735 | 0.345 (0.329 - 0.362 ) | 8602/6120223 | 1.41 (1.38 - 1.44 ) |
| DCM | missense pLOF FAF | G- | 79 | 0.0429 (0.034 - 0.0535 ) | 326/2238615 | 0.146 (0.13 - 0.162 ) |
|  |  | G+ | 23 | 0.138 (0.0875 - 0.207 ) | 61/201790 | 0.302 (0.231 - 0.388 ) |
|  |  | Unknown | 229 | 0.0759 (0.0664 - 0.0864 ) | 697/3729230 | 0.187 (0.173 - 0.201 ) |
|  | InterVar FAF | G- | 98 | 0.0493 (0.04 - 0.06 ) | 369/2419884 | 0.152 (0.137 - 0.169 ) |
|  |  | G+ | 4 | 0.236 (0.0642 - 0.602 ) | 18/20520 | 0.877 (0.52 - 1.39 ) |
|  |  | Unknown | 229 | 0.0759 (0.0664 - 0.0864 ) | 697/3729230 | 0.187 (0.173 - 0.201 ) |
|  | InterVar FAF ClinVar 2* | G- | 94 | 0.0474 (0.0383 - 0.058 ) | 368/2412675 | 0.153 (0.137 - 0.169 ) |
|  |  | G+ | 8 | 0.348 (0.15 - 0.684 ) | 19/27729 | 0.685 (0.413 - 1.07 ) |
|  |  | Unknown | 229 | 0.0759 (0.0664 - 0.0864 ) | 697/3729230 | 0.187 (0.173 - 0.201 ) |
|  | Total | NA | 331 | 0.0659 (0.059 - 0.0734 ) | 1084/6169635 | 0.176 (0.165 - 0.186 ) |

For each filtering strategy and genotype status, the prevalence (%) of the listed phenotypes at study entry and incidence per 1000 person-years of follow up is shown. N = number of participants with phenotype of interest at study entry; CI = 95% confidence interval; DCM = dilated cardiomyopathy; CIED = cardiac implantable electronic device (including single- and dual-chamber permanent pacemaker, implantable cardioverter defibrillator, cardiac resynchronization therapy).

**Table S9. Prevalence and incidence of DCM and DCM-associated features in study population.**

|  |  |  | Prevalence | | Incidence | |
| --- | --- | --- | --- | --- | --- | --- |
|  | Filter | Genotype | N | Prevalence (CI) [%] | Events/Person-Time | Incidence (CI) |
| AF | missense pLOF FAF | G- | 170 | 0.988 (0.846 - 1.15 ) | 532/211054 | 2.52 (2.31 - 2.74 ) |
|  |  | G+ | 12 | 0.82 (0.425 - 1.43 ) | 50/17954 | 2.78 (2.07 - 3.67 ) |
|  | InterVar FAF | G- | 181 | 0.978 (0.841 - 1.13 ) | 573/227140 | 2.52 (2.32 - 2.74 ) |
|  |  | G+ | 1 | 0.649 (0.0164 - 3.56 ) | 9/1868 | 4.82 (2.2 - 9.15 ) |
|  | InterVar FAF ClinVar 2* | G- | 180 | 0.975 (0.839 - 1.13 ) | 570/226428 | 2.52 (2.31 - 2.73 ) |
|  |  | G+ | 2 | 0.943 (0.114 - 3.37 ) | 12/2580 | 4.65 (2.4 - 8.12 ) |
|  | Total | NA | 182 | 0.975 (0.839 - 1.13 ) | 582/229008 | 2.54 (2.34 - 2.76 ) |
| Bradyarrhythmia | missense pLOF FAF | G- | 107 | 0.622 (0.51 - 0.751 ) | 200/213715 | 0.936 (0.811 - 1.07 ) |
|  |  | G+ | 12 | 0.82 (0.425 - 1.43 ) | 15/18083 | 0.829 (0.464 - 1.37 ) |
|  | InterVar FAF | G- | 118 | 0.637 (0.528 - 0.763 ) | 214/229895 | 0.931 (0.81 - 1.06 ) |
|  |  | G+ | 1 | 0.649 (0.0164 - 3.56 ) | 1/1904 | 0.525 (0.0133 - 2.93 ) |
|  | InterVar FAF ClinVar 2* | G- | 117 | 0.634 (0.525 - 0.759 ) | 214/229174 | 0.934 (0.813 - 1.07 ) |
|  |  | G+ | 2 | 0.943 (0.114 - 3.37 ) | 1/2624 | 0.381 (0.00965 - 2.12 ) |
|  | Total | NA | 119 | 0.638 (0.528 - 0.762 ) | 215/231798 | 0.928 (0.808 - 1.06 ) |
| Conduction Defect | missense pLOF FAF | G- | 167 | 0.971 (0.83 - 1.13 ) | 99/213293 | 0.464 (0.377 - 0.565 ) |
|  |  | G+ | 12 | 0.82 (0.425 - 1.43 ) | 9/18098 | 0.497 (0.227 - 0.944 ) |
|  | InterVar FAF | G- | 178 | 0.962 (0.826 - 1.11 ) | 107/229489 | 0.466 (0.382 - 0.563 ) |
|  |  | G+ | 1 | 0.649 (0.0164 - 3.56 ) | 1/1903 | 0.526 (0.0133 - 2.93 ) |
|  | InterVar FAF ClinVar 2* | G- | 178 | 0.965 (0.829 - 1.12 ) | 106/228758 | 0.463 (0.379 - 0.56 ) |
|  |  | G+ | 1 | 0.472 (0.0119 - 2.6 ) | 2/2633 | 0.759 (0.092 - 2.74 ) |
|  | Total | NA | 179 | 0.959 (0.824 - 1.11 ) | 108/231392 | 0.467 (0.383 - 0.564 ) |
| Sick sinus sindrome | missense pLOF FAF | G- | 16 | 0.093 (0.0532 - 0.151 ) | 1/215561 | 0.00464 (0.000117 - 0.0258 ) |
|  |  | G+ | 2 | 0.137 (0.0166 - 0.493 ) | 0/18254 | 0 (0 - 0.202 ) |
|  | InterVar FAF | G- | 18 | 0.0972 (0.0576 - 0.154 ) | 1/231899 | 0.00431 (0.000109 - 0.024 ) |
|  |  | G+ | 0 | 0 (0 - 2.37 ) | 0/1916 | 0 (0 - 1.93 ) |
|  | InterVar FAF ClinVar 2* | G- | 18 | 0.0975 (0.0578 - 0.154 ) | 1/231164 | 0.00433 (0.00011 - 0.0241 ) |
|  |  | G+ | 0 | 0 (0 - 1.72 ) | 0/2651 | 0 (0 - 1.39 ) |
|  | Total | NA | 18 | 0.0964 (0.0572 - 0.152 ) | 1/233815 | 0.00428 (0.000108 - 0.0238 ) |
| Pre-excitation syndrome | missense pLOF FAF | G- | 17 | 0.0988 (0.0576 - 0.158 ) | 11/215476 | 0.051 (0.0255 - 0.0913 ) |
|  |  | G+ | 1 | 0.0684 (0.00173 - 0.38 ) | 1/18262 | 0.0548 (0.00139 - 0.305 ) |
|  | InterVar FAF | G- | 18 | 0.0972 (0.0576 - 0.154 ) | 12/231822 | 0.0518 (0.0267 - 0.0904 ) |
|  |  | G+ | 0 | 0 (0 - 2.37 ) | 0/1916 | 0 (0 - 1.93 ) |
|  | InterVar FAF ClinVar 2* | G- | 17 | 0.0921 (0.0537 - 0.147 ) | 11/231101 | 0.0476 (0.0238 - 0.0852 ) |
|  |  | G+ | 1 | 0.472 (0.0119 - 2.6 ) | 1/2637 | 0.379 (0.0096 - 2.11 ) |
|  | Total | NA | 18 | 0.0964 (0.0572 - 0.152 ) | 12/233738 | 0.0513 (0.0265 - 0.0897 ) |
| Ventricular arrhythmia | missense pLOF FAF | G- | 9 | 0.0523 (0.0239 - 0.0993 ) | 53/215454 | 0.246 (0.184 - 0.322 ) |
|  |  | G+ | 0 | 0 (0 - 0.252 ) | 7/18261 | 0.383 (0.154 - 0.79 ) |
|  | InterVar FAF | G- | 9 | 0.0486 (0.0222 - 0.0923 ) | 59/231802 | 0.255 (0.194 - 0.328 ) |
|  |  | G+ | 0 | 0 (0 - 2.37 ) | 1/1913 | 0.523 (0.0132 - 2.91 ) |
|  | InterVar FAF ClinVar 2* | G- | 9 | 0.0488 (0.0223 - 0.0926 ) | 58/231071 | 0.251 (0.191 - 0.324 ) |
|  |  | G+ | 0 | 0 (0 - 1.72 ) | 2/2644 | 0.757 (0.0916 - 2.73 ) |
|  | Total | NA | 9 | 0.0482 (0.0221 - 0.0915 ) | 60/233715 | 0.257 (0.196 - 0.33 ) |
| Heart failure | missense pLOF FAF | G- | 27 | 0.157 (0.103 - 0.228 ) | 183/214723 | 0.852 (0.733 - 0.985 ) |
|  |  | G+ | 7 | 0.478 (0.193 - 0.983 ) | 23/18095 | 1.27 (0.806 - 1.91 ) |
|  | InterVar FAF | G- | 32 | 0.173 (0.118 - 0.244 ) | 201/230950 | 0.87 (0.754 - 0.999 ) |
|  |  | G+ | 2 | 1.3 (0.158 - 4.61 ) | 5/1868 | 2.68 (0.869 - 6.25 ) |
|  | InterVar FAF ClinVar 2* | G- | 31 | 0.168 (0.114 - 0.238 ) | 199/230234 | 0.864 (0.748 - 0.993 ) |
|  |  | G+ | 3 | 1.42 (0.293 - 4.08 ) | 7/2584 | 2.71 (1.09 - 5.58 ) |
|  | Total | NA | 34 | 0.182 (0.126 - 0.254 ) | 206/232818 | 0.885 (0.768 - 1.01 ) |
| Stroke | missense pLOF FAF | G- | 118 | 0.686 (0.568 - 0.821 ) | 183/213349 | 0.858 (0.738 - 0.991 ) |
|  |  | G+ | 13 | 0.889 (0.474 - 1.51 ) | 16/18031 | 0.887 (0.507 - 1.44 ) |
|  | InterVar FAF | G- | 130 | 0.702 (0.587 - 0.833 ) | 199/229477 | 0.867 (0.751 - 0.996 ) |
|  |  | G+ | 1 | 0.649 (0.0164 - 3.56 ) | 0/1903 | 0 (0 - 1.94 ) |
|  | InterVar FAF ClinVar 2* | G- | 130 | 0.704 (0.589 - 0.836 ) | 198/228745 | 0.866 (0.749 - 0.995 ) |
|  |  | G+ | 1 | 0.472 (0.0119 - 2.6 ) | 1/2635 | 0.379 (0.00961 - 2.11 ) |
|  | Total | NA | 131 | 0.702 (0.587 - 0.832 ) | 199/231380 | 0.86 (0.745 - 0.988 ) |
| CIED | missense pLOF FAF | G- | 10 | 0.0581 (0.0279 - 0.107 ) | 123/215269 | 0.571 (0.475 - 0.682 ) |
|  |  | G+ | 0 | 0 (0 - 0.252 ) | 15/18227 | 0.823 (0.461 - 1.36 ) |
|  | InterVar FAF | G- | 10 | 0.054 (0.0259 - 0.0993 ) | 137/231582 | 0.592 (0.497 - 0.699 ) |
|  |  | G+ | 0 | 0 (0 - 2.37 ) | 1/1914 | 0.523 (0.0132 - 2.91 ) |
|  | InterVar FAF ClinVar 2* | G- | 10 | 0.0542 (0.026 - 0.0996 ) | 136/230850 | 0.589 (0.494 - 0.697 ) |
|  |  | G+ | 0 | 0 (0 - 1.72 ) | 2/2646 | 0.756 (0.0916 - 2.73 ) |
|  | Total | NA | 10 | 0.0536 (0.0257 - 0.0985 ) | 138/233496 | 0.591 (0.497 - 0.698 ) |
| DCM | missense pLOF FAF | G- | 4 | 0.0233 (0.00634 - 0.0595 ) | 10/215674 | 0.0464 (0.0222 - 0.0853 ) |
|  |  | G+ | 1 | 0.0684 (0.00173 - 0.38 ) | 4/18258 | 0.219 (0.0597 - 0.561 ) |
|  | InterVar FAF | G- | 5 | 0.027 (0.00877 - 0.063 ) | 12/232020 | 0.0517 (0.0267 - 0.0903 ) |
|  |  | G+ | 0 | 0 (0 - 2.37 ) | 2/1912 | 1.05 (0.127 - 3.78 ) |
|  | InterVar FAF ClinVar 2* | G- | 5 | 0.0271 (0.0088 - 0.0632 ) | 11/231288 | 0.0476 (0.0237 - 0.0851 ) |
|  |  | G+ | 0 | 0 (0 - 1.72 ) | 3/2644 | 1.13 (0.234 - 3.32 ) |
|  | Total | NA | 5 | 0.0268 (0.0087 - 0.0625 ) | 14/233932 | 0.0598 (0.0327 - 0.1 ) |

For each filtering strategy and genotype status, the prevalence (%) of the listed phenotypes at study entry and Incidence per 1000 person-years of follow up is shown. N = number of participants with phenotype of interest at study entry; CI = 95% confidence interval; DCM = dilated cardiomyopathy; CIED = cardiac implantable electronic device (including single- and dual-chamber permanent pacemaker, implantable cardioverter defibrillator, cardiac resynchronization therapy).

**Table S10.** STROBE Statement—Checklist of items that should be included in reports of ***cohort studies***

|  | Item No | Recommendation | Page Number |
| --- | --- | --- | --- |
| **Title and abstract** | 1 | (*a*) Indicate the study’s design with a commonly used term in the title or the abstract | 2 |
|  |  | (*b*) Provide in the abstract an informative and balanced summary of what was done and what was found | 2 |
| Introduction | | |  |
| Background/rationale | 2 | Explain the scientific background and rationale for the investigation being reported | 4 |
| Objectives | 3 | State specific objectives, including any prespecified hypotheses | 4 |
| Methods | | |  |
| Study design | 4 | Present key elements of study design early in the paper | 6-9 |
| Setting | 5 | Describe the setting, locations, and relevant dates, including periods of recruitment, exposure, follow-up, and data collection | 6-9 |
| Participants | 6 | (*a*) Give the eligibility criteria, and the sources and methods of selection of participants. Describe methods of follow-up | 6-9, Figure 1 |
|  |  | (*b*) For matched studies, give matching criteria and number of exposed and unexposed | NA |
| Variables | 7 | Clearly define all outcomes, exposures, predictors, potential confounders, and effect modifiers. Give diagnostic criteria, if applicable | 6-9, Table S9 |
| Data sources/ measurement | 8* | For each variable of interest, give sources of data and details of methods of assessment (measurement). Describe comparability of assessment methods if there is more than one group | 6-9 |
| Bias | 9 | Describe any efforts to address potential sources of bias | 6-9 |
| Study size | 10 | Explain how the study size was arrived at | 6-9, Figure 1 |
| Quantitative variables | 11 | Explain how quantitative variables were handled in the analyses. If applicable, describe which groupings were chosen and why | 6-9 |
| Statistical methods | 12 | (*a*) Describe all statistical methods, including those used to control for confounding | 9, supplementary methods |
|  |  | (*b*) Describe any methods used to examine subgroups and interactions | 6-9 |
|  |  | (*c*) Explain how missing data were addressed | NA |
|  |  | (*d*) If applicable, explain how loss to follow-up was addressed | NA |
|  |  | (*e*) Describe any sensitivity analyses | NA |
| Results | | |  |
| Participants | 13* | (a) Report numbers of individuals at each stage of study—eg numbers potentially eligible, examined for eligibility, confirmed eligible, included in the study, completing follow-up, and analysed | 10, Figure 1 |
|  |  | (b) Give reasons for non-participation at each stage | Figure 1 |
|  |  | (c) Consider use of a flow diagram | Figure 1 |
| Descriptive data | 14* | (a) Give characteristics of study participants (eg demographic, clinical, social) and information on exposures and potential confounders | 10, Table 1 |
|  |  | (b) Indicate number of participants with missing data for each variable of interest | NA |
|  |  | (c) Summarise follow-up time (eg, average and total amount) | Table S4 |
| Outcome data | 15* | Report numbers of outcome events or summary measures over time | Figure S2 |
| Main results | 16 | (*a*) Give unadjusted estimates and, if applicable, confounder-adjusted estimates and their precision (eg, 95% confidence interval). Make clear which confounders were adjusted for and why they were included | 10-11, Table S4 |
|  |  | (*b*) Report category boundaries when continuous variables were categorized | NA |
|  |  | (*c*) If relevant, consider translating estimates of relative risk into absolute risk for a meaningful time period | NA |
| Other analyses | 17 | Report other analyses done—eg analyses of subgroups and interactions, and sensitivity analyses | NA |
| Discussion | | |  |
| Key results | 18 | Summarise key results with reference to study objectives | 11 |
| Limitations | 19 | Discuss limitations of the study, taking into account sources of potential bias or imprecision. Discuss both direction and magnitude of any potential bias | 14 |
| Interpretation | 20 | Give a cautious overall interpretation of results considering objectives, limitations, multiplicity of analyses, results from similar studies, and other relevant evidence | 14-15 |
| Generalisability | 21 | Discuss the generalisability (external validity) of the study results | 12-15 |
| Other information | | |  |
| Funding | 22 | Give the source of funding and the role of the funders for the present study and, if applicable, for the original study on which the present article is based | 16 |

*Give information separately for exposed and unexposed groups.

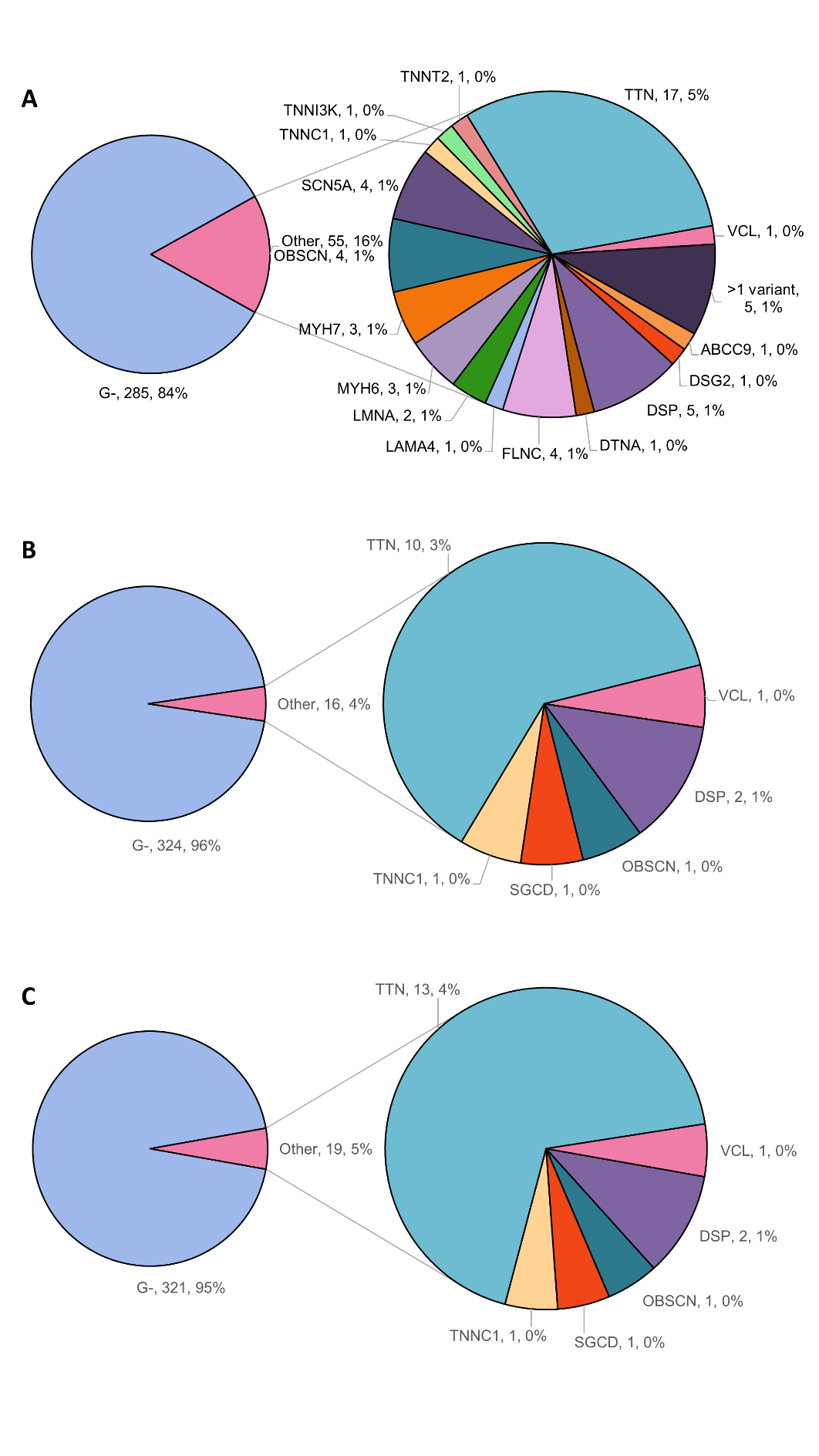

**Figure S1. Genetic yield of clinically diagnosed dilated cardiomyopathy (DCM) by the three variant filtering strategies.**

A. missense pLOF FAF filtering strategy; B. InterVar FAF filtering strategy; C. InterVar FAF Clinvar variant filtering strategy. For participants with clinically diagnosed DCM, the pie chart shows the percentage and number with a putative pathogenic variant in a DCM-related gene, or if no variant identified (G–).

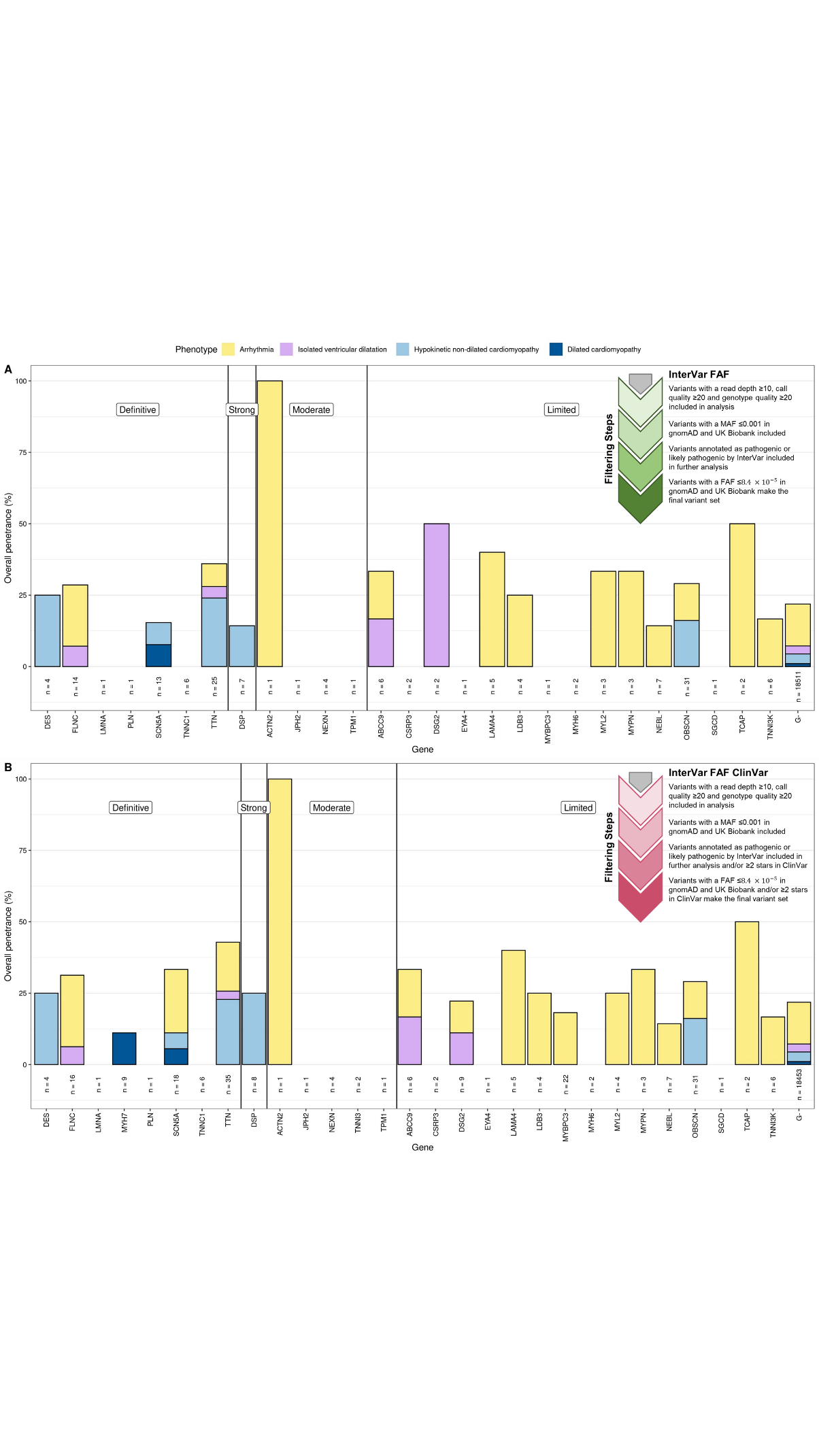
**Figure S2. Clinical and subclinical penetrance of DCM-associated genes for secondary variant filtering strategies.**

For each DCM-associated gene, the height of the bar indicates the percentage of putative pathogenic variant mutation carriers, by the relevant filtering strategy, with the specified phenotypes. Results are show for InterVar FAF (upper panel), and InterVar FAF Clinvar (lower panel). Total number of participants with a putative pathogenic variant for each DCM-associated gene is indicated below the bar. The phenotype prevalence in those without a putative pathogenic variant is show on the far right (labelled “G-“). Genes are categorized according to the strength of evidence determined by the ClinGen DCM Gene Expert Curation Panel and ordered alphabetically within each category.

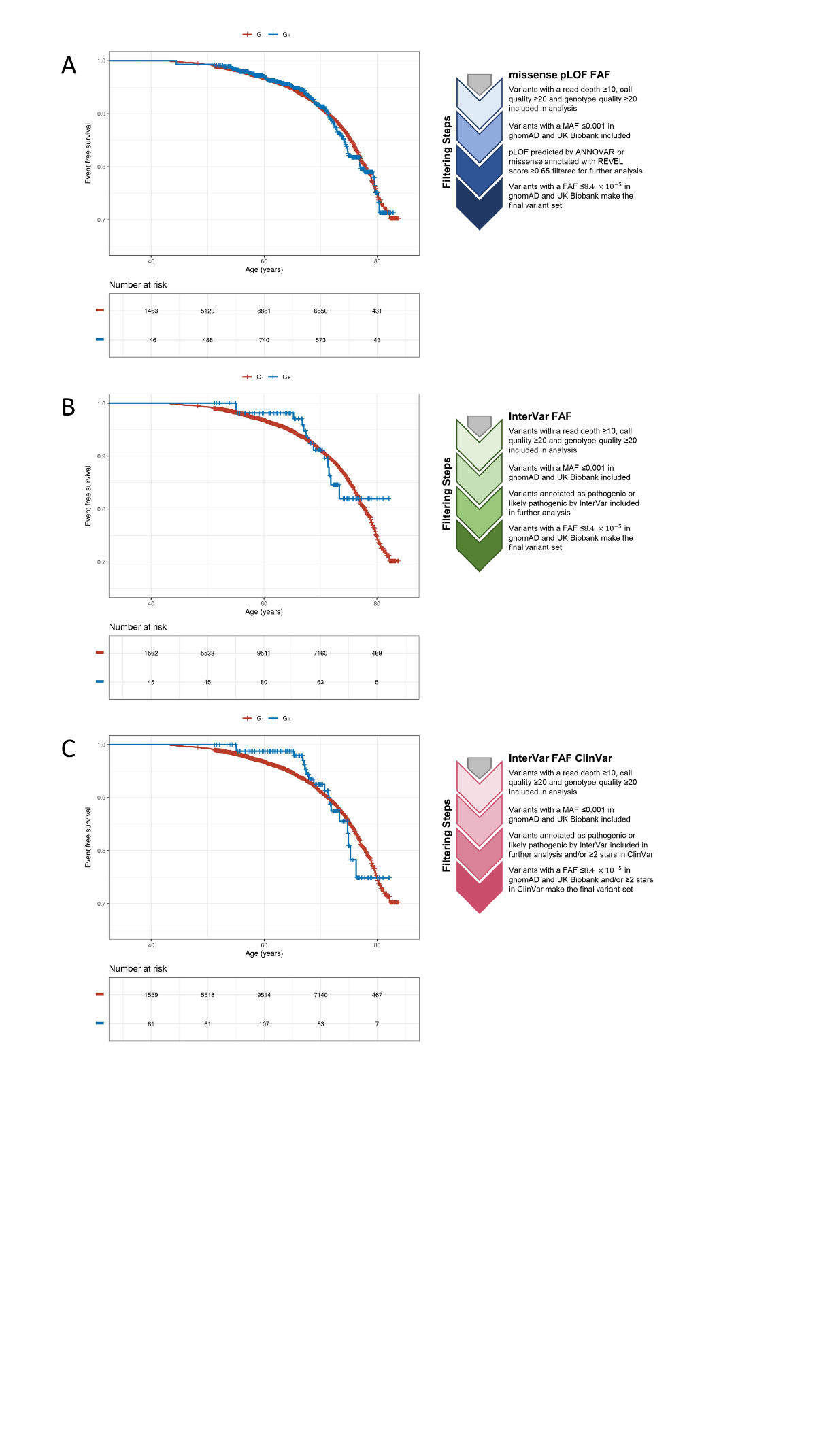
**Figure S3.** Kaplan-Meier plots showing event-free survival for genotype-positive versus genotype-negative patients for each of the three filtering strategies: (A) missense pLOF FAF, (B) InterVar FAF, and (C) InterVar FAF ClinVar. Event free survival was defined as survival without developing one of heart failure, stroke, arrhythmia, required CIED, or death.

CIED = cardiac implantable electronic device (including single- and dual-chamber permanent pacemaker, implantable cardioverter defibrillator, cardiac resynchronization therapy)
